# Assessing the knowledge, attitudes and practices of healthcare staff and students regarding disposal of unwanted medications: A systematic review

**DOI:** 10.1101/2024.11.17.24317468

**Authors:** Janeme Lam, Dayana El Nsouli, E Lyn Lee, Tawfiq Alqeisi, Ros Kane, Ian McGonagle, Despina Laparidou, David Nelson, Keivan Armani

## Abstract

**Objectives:** We sought to review studies that examine healthcare professionals’ and students’ knowledge, attitudes, and practices (KAP) regarding medication disposal. We also explore recommendations and barriers related to appropriate medication disposal.

**Design:** A systematic review was conducted that adhered to Preferred Reporting Items for Systematic reviews and Meta-Analyses (PRISMA).

**Data sources:** MEDLINE, Embase, CINAHL, Web of Science, PsycINFO and Google Scholar were searched up to 23^rd^ February 2024.

**Study eligibility criteria and setting:** Qualitative, quantitative, and mixed-method primary research studies. There was no limitation on the publication date, geographical locations, or the study settings.

**Participants:** Pharmacists, doctors, nurses, and students from these respective professional groups in any country.

**Primary outcome measures:** The levels of healthcare staff and students’ knowledge, attitudes, and practices about disposal of unwanted medications.

**Date extraction and synthesis:** Data extraction was conducted by four of the researchers independently. The study details were categorised into three main domains, i.e., knowledge, attitude, and practice using the KAP model. Other relevant information was also extracted, and synthesised in overall themes, such as challenges and recommendations.

**Results:** 37 studies from 18 countries (Asia n=21; USA n=7; Africa n=5; and EU n=2; South America n=2) were included. 86.5% (n=32) investigated participants’ knowledge of medication disposal. Although there was a good level of awareness about the environmental impacts, there were significant gaps in knowledge regarding correct disposal methods, available services, guidelines, and training. 30 studies explored participants’ attitudes toward medication disposal. There was a generally positive attitude towards the need for environmentally safe disposal practices. 35 studies evaluated participants’ practices in relation to medication disposal. Although there was generally a positive attitude and some understanding of appropriate disposal methods, the majority of the participants did not follow the practice guidelines, especially outside healthcare settings.

**Discussions and Conclusions:** While healthcare staff and students have fair knowledge and positive attitudes toward medicine disposal, their actual practices are lacking. One significant challenge identified is the limited awareness about proper disposal methods coupled with a lack of established services or guidelines. Even in cases where take-back programs are available, they often face issues with accessibility. To tackle these challenges, it is suggested that governmental bodies should establish and enforce clear policies on medication disposal while also expanding educational initiatives to increase understanding among professionals and students. Furthermore, improving access to take-back programs is crucial for ensuring safe medication disposal and minimising potential environmental and health hazards.

**PROSPERO registration number:** CRD42024503162.

**Strengths and limitations of this study:**

- The review used the theoretical framework (KAP model) to effectively structure the literature search and data organisation, ensuring a comprehensive focus on studies and consistent data collection and analysis.
- This review covered all three components of the KAP model that is knowledge, attitude and practice, resulting in a more robust data synthesis with increased generalisability.
- The review encompassed a considerable number of studies (n=37) conducted in 18 different countries across five continents. The diverse range of populations and settings enhances the broad relevance of the review’s results.
- The review only included studies in English language.

## Introduction

Pharmaceutical waste is any waste which contains medicinal drugs that are expired, unused, contaminated, damaged, or no longer needed.[1,2] Inappropriate handling of pharmaceutical waste may lead to serious public health consequences and negative impact on the environment.[3] It has been shown that approximately 10% of pharmaceutical products have a significant potential environmental risk, especially active medications.[4] In addition to the environmental impact, improper disposal of medicines can lead to risk of misuse, drug diversion, accidental ingestion, especially in households with children, younger adults, or pets.[5] Therefore, building a comprehensive understanding of the impact of pharmaceutical waste on the environment and public health is essential for all individuals involved in healthcare.

The proper disposal of unwanted medications is a significant global issue. Several developed nations have implemented initiatives to address the disposal of unwanted medicines. For example, Australia and Canada have launched the National Return and Disposal of Unwanted Medicines Project with full support from their governments and pharmaceutical industries.[6] Similarly, drug take-back programs are widely established in Sweden.[7] In the United Kingdom, community pharmacy contractors are obliged to accept back unwanted medicines from patients.[8,9] However, low- and middle-income countries still lack widespread advocacy for safe disposal practices of unused medicines by the public.[10]

The World Health Organization (WHO) has issued recommendations for the proper disposal of pharmaceuticals to minimise the potential impact on human health and the environment. These guidelines are intended for adoption in areas at national or regional levels that may lack comprehensive regulations or frameworks for pharmaceutical disposal.[11] The recommendation suggests that pharmaceuticals should be disposed of through high-temperature incineration, preferably at temperatures above 1,200°C. However, other methods can also be utilised for proper disposal as not all countries have access to such facilities.

A systematic review was undertaken to evaluate the public’s knowledge, attitude and practice concerning the disposal of household medication waste.[12] The majority of the included studies indicated that knowledge about safe medication disposal was generally limited to inadequate. Despite this, there was a positive attitude towards proper disposal. However, actual disposal practices were found to be insufficient, likely due to a lack of accessible disposal facilities, information, and guidance from healthcare professionals.

Healthcare professionals can play a pivotal role in educating the public about safe disposal of medications. Therefore, understanding their knowledge, attitudes, and practices (KAP) regarding medication disposal is essential. This systematic review identified and analysed studies that examine healthcare professionals’ and students’ KAP regarding disposal of unwanted medicines. It also explored recommendations and obstacles related to appropriate medication disposal.

The concept of knowledge, attitude, and practice (KAP) was first introduced by Hochbaum[13] and utilised to evaluate family planning in the 1950s.[13,14] According to the KAP model, practice is influenced by attitude and knowledge. It suggests that individuals with a strongly positive attitude toward a certain behaviour and extensive knowledge will engage in effective practices. This model allows us to understand not only individual beliefs and knowledge but also how these attitudes impact behaviour.[14] Synthesising this evidence is important because it provides a comprehensive understanding of the current state of the knowledge, attitude and practice of medicines disposal, highlighting areas that require improvement. Moreover, it enables the development of targeted interventions and policies to enhance medication disposal practices, ultimately promoting environmental safety and public health.

## Methods

This systematic review is reported in accordance with the Preferred Reporting Items for Systematic review and Meta-Analysis’ guidelines (PRISMA) (Appendix 1). The study protocol is registered with International Prospective Register of Systematic Reviews (PROSPERO) (http://www.crd.york.ac.uk/PROSPERO) [Protocol registration number: CRD42024503162] (see Supplementary material appendix 2).

### Study characteristics

Authors included qualitative, quantitative, and mixed-method primary research studies and only studies published in English. Full details for the inclusion and exclusion criteria are shown in Table 1. There was no limitation on the publication date, geographical locations, or the settings of the studies.

**Table 1.**
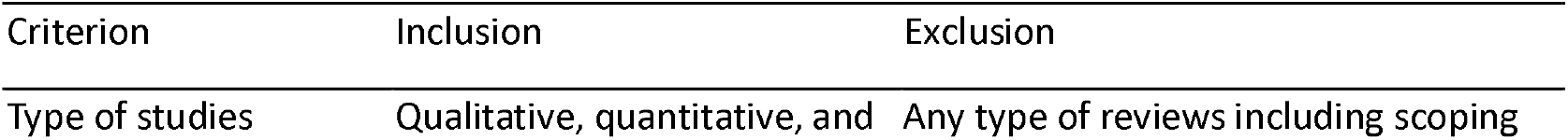
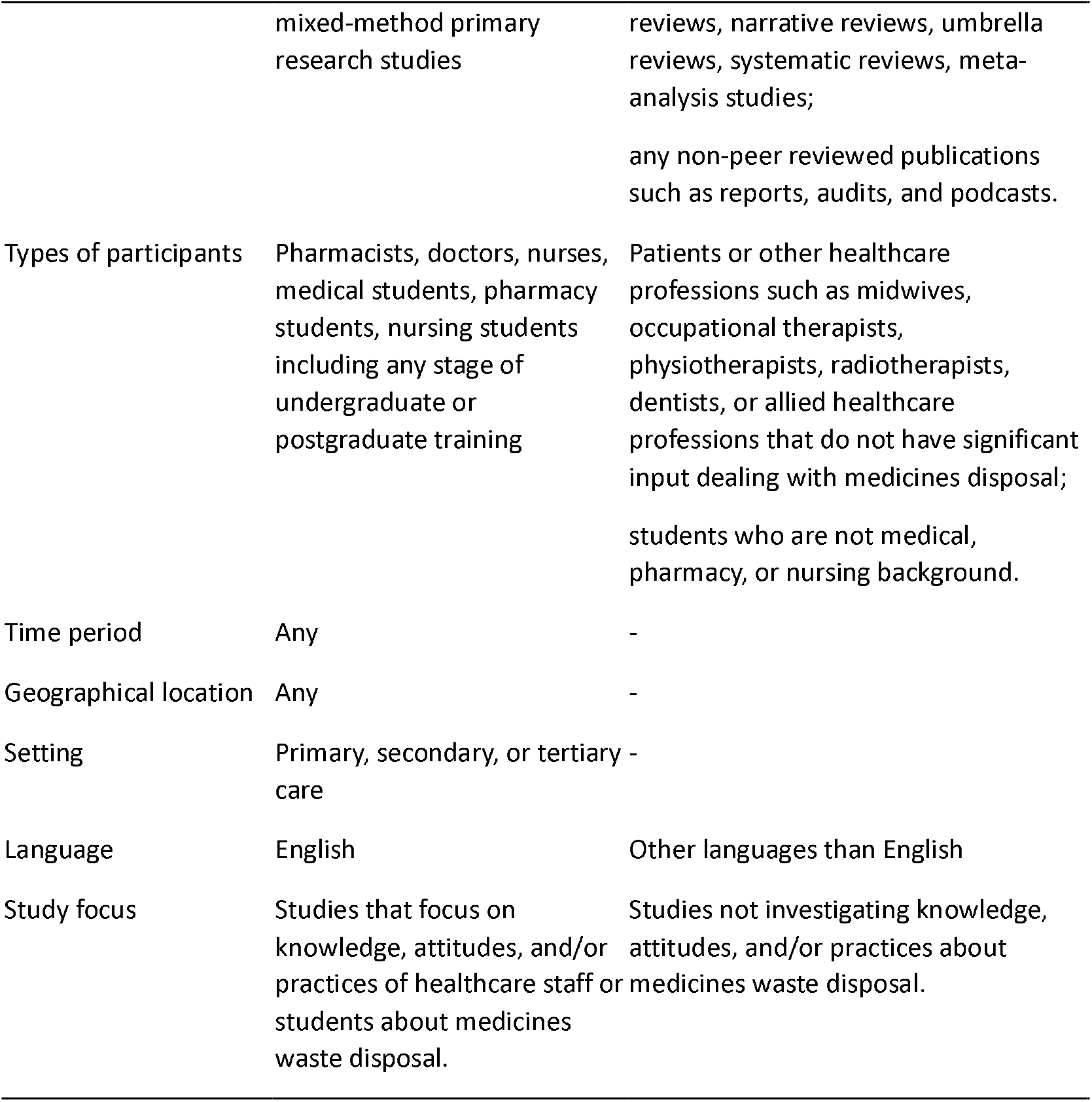
Inclusion and Exclusion criteria.

Participants included pharmacists, doctors, nurses, and both undergraduate and postgraduate medical, pharmacy, and nursing students due to their significant involvement with medication and its disposal in healthcare settings.

The main outcome of this review was to measure the levels of the healthcare staff and students’ knowledge, attitudes, and/or practices about medicines waste disposal at any healthcare settings. Studies that mentioned either of the three components of the KAP or all of them were included. While the studies primarily focused on the knowledge, attitude and practice of medicines disposal, many of them provided valuable insights into the challenges faced in implementing proper disposal practices and suggested strategies for improvement. These insights were categorised and analysed to identify common themes and actionable recommendations.

## Search methods for identification of studies

### Electronic searches

The lead author (JL) developed a search strategy in collaboration with two medical librarians and with support from other members of the review team (DEN and KA). The syntax that was used to search the databases incorporated key words, phrases and medical subject headings for three main concepts: (1) pharmacists, doctors, nurses, pharmacy students, medical students, nursing students, (2) knowledge, attitude, practice, and (3) medicine disposal (see Appendix 3 for details). The search strategy was conducted on multiple bibliographic databases up to 23^rd^ February 2024. The databases searched were as follows: MEDLINE, Embase, CINAHL, Web of Science, PsycINFO. These searches were supplemented with internet searches (i.e. Google Scholar). A summary of the contents of each database can be found in Table 2.

Searches were conducted without limits to location or publication date. Supplementary material appendix 3 provides an overview of the search terms and results for the various databases.

All references were imported into Covidence™, and duplicates removed prior to title and abstract screening. Additionally, the abstracts were uploaded into Covidence™. The platform was also used for full-text screening, data extraction, and quality assessment.

Two researchers (JL and DEN) independently screened all titles and abstracts for eligibility and subsequently reviewed the full-text articles for inclusion. Discrepancies were resolved through discussions or with the support of a third researcher (KA).

## Data collection and analysis

### Data extraction and management

Authors created a bespoke data extraction form by adjusting the pre-determined Covidence™ data extraction template. See supplementary material appendix 4. The extraction sheet captured the following information:

- Study characteristics: study type, setting, geographic location, study population, number of respondents, data collection period.
- Results of included studies with respect to outcomes of knowledge, attitude, practice, legislation, challenges, and recommendations.

Data extraction involved four researchers (JL, DEN, ELL, TA), with data being independently extracted by two randomly assigned researchers. Any discrepancies for the data extraction amongst the researchers were resolved by KA, who was not involved with the initial data extraction.

### Quality assessment

Quality and risk of bias were assessed using the Joanna Briggs Institute’s (JBI) critical appraisal tools. For analytical cross-sectional studies, five questions were included after excluding three irrelevant ones (Q3, Q5, Q6) related to exposure and confounding factors. Each question required a yes, no, or unclear answer.

### Data synthesis

The study details were categorised into three main headings i.e., knowledge, attitude, and practice using the KAP model.[14] This categorisation allowed a systematic and robust operationalisation of the findings to answer the research objectives. Other relevant information was also extracted, and synthesised in overall themes, such as challenges and recommendations. These recommendations and barriers were extracted from the discussion and conclusion sections of the studies reviewed. For each sub-theme, the level of knowledge was scored as poor (<50%), fair (50%-75%) or satisfactory (≥75%). The attitude level for each sub-theme was assessed as either negative (<50%) or positive (≥50%), based on the percentages obtained from the included studies. These categorisations were selected based on precedent in the literature to provide a clear and standardised method for assessing the levels of knowledge and attitudes.[12,15] The themes emerged through an iterative process of reading the full articles, familiarising with the data, grouping results into overarching themes. Practices were classified into two main practice domains and further into two subdomains. The practice domains were: 1) medicine disposal outside healthcare settings; 2) medicine disposal within healthcare settings.

## Results

A total of 733 articles were identified across six databases, i.e., MEDLINE (n = 100), Embase (n = 100), CINAHL (n=94), Web of Science (n = 200), Google Scholar (n=221), and PsycINFO (n=18). Articles were exported into Covidence™ for further processing. Following the removal of duplicates, 496 studies were screened by title and abstract. After this, a total of 75 articles were then screened by full-text against the eligibility criteria. The full-text screening eliminated 31 articles because of wrong outcome i.e. not assessing KAP (n = 18), wrong study design (n = 5), wrong population (n=5), wrong setting (n=2), and not in English (n = 1) and thus, 37 studies were included in the final review upon which results are reported (see Figure 1).

**Figure 1.**
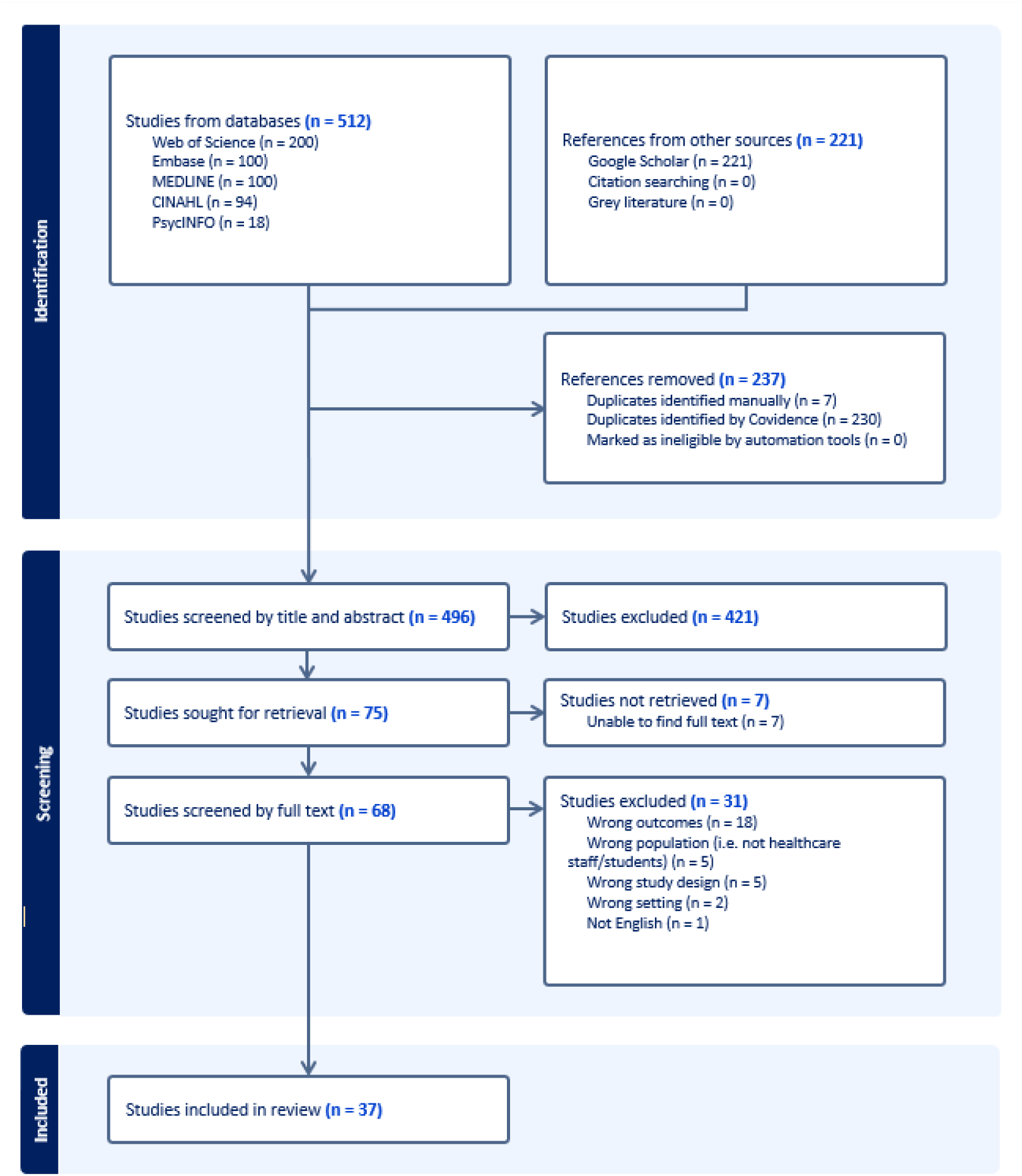
PRISMA diagram of the systematic review

Most studies covered all three domains (n=27)[16–42], while 6 focused on knowledge and practices[43–48], and 4 on attitudes and practices[49–52]. All studies explored the challenges and recommendations for implementing appropriate medicine disposal practices.

## Types of studies, location and year

All studies were published from 2009 to 20^th^ December 2023. The majority of the studies were survey-based quantitative studies (n=34), and a smaller number employing mixed methods studies (n=3)[50–52]. Two of the studies assessed knowledge and practice pre- and post-educational interventions.[29,48] Studies were conducted in 18 different countries, with the majority originating from Asia (n=21)[16–20,22,23,30,31,34,36–39,41,44,45,47–50], seven from the USA[21,25,29,32,35,42,51], five from Africa[26,27,43,46,52], two from the EU[24,40] and two from South America[28,33] (see Figure 2). The countries included were Bangladesh, Brazil, Ethiopia, India, Indonesia, Iraq, Kosovo, Kuwait, Malaysia, Nepal, Nigeria, Palestine, Romania, Saudi Arabia, South Africa, Trinidad and Tobago, United Arab Emirates, and the United States.

**Figure 2.**
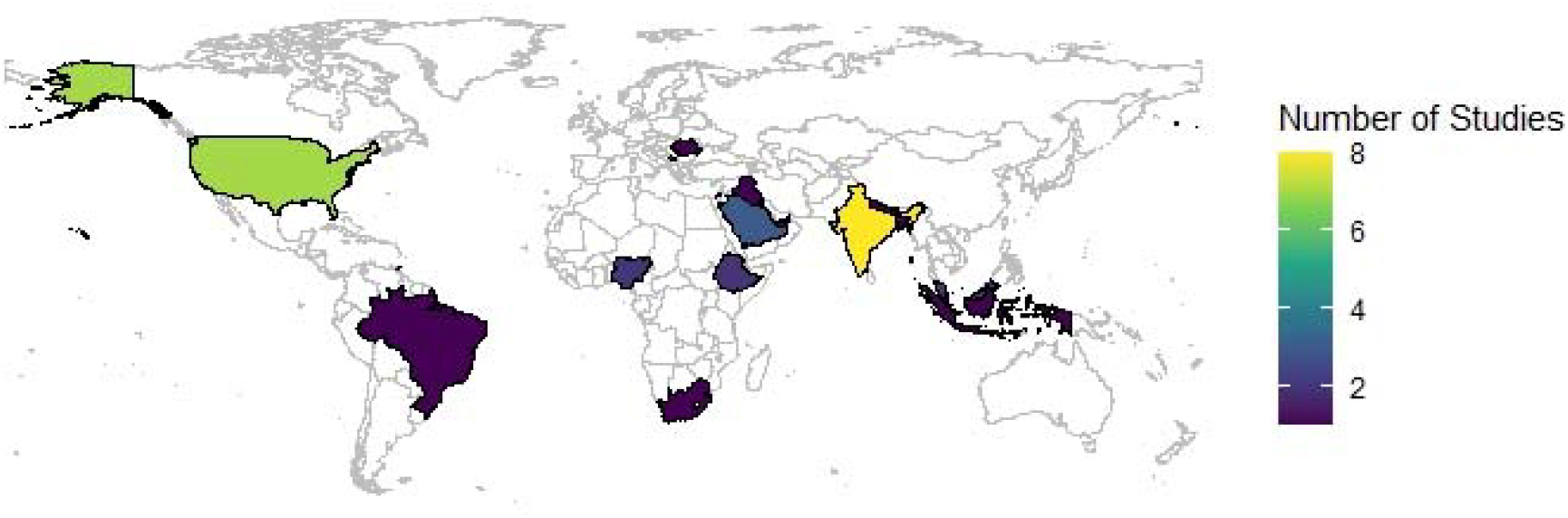
Country of origin of the articles included in this review

## Studies’ participants

Overall, 9952 participants were included in 37 studies. Nearly three-quarters of the studies (n=26) focused on healthcare professionals; predominantly pharmacists (n=18)[16–19,22,28–31,33– 35,37,42,44,50–52], nurses (n=1)[32], and combined participants of pharmacists, nurses, and doctors (n=4)[20,36,46,49]. Less than a third of the studies (n=10) assessed healthcare students, specifically pharmacy students (n=3)[25,26,38], medical students (n=3)[39,41,45], and multiple professions such as pharmacy, nursing, and/or medical students (n=4)[21,23,40,43]. One study included both healthcare staff (doctors and nurses) and students (pharmacy and medical students).[47]

## Knowledge domain

Among the included studies, 86.5% (n=32) investigated participants’ knowledge of medication disposal (see Figure 3).[16–44,46–48] Out of 20 studies that examined the knowledge of environmental impact, 95% reported fair to satisfactory levels.[16,18– 20,22,23,26,28,29,31,33,34,36,38,40,41,43,47,48] In assessing awareness of service availability, a quarter out of the 8 studies reported poor knowledge levels.[31,38] As for the 10 studies measuring awareness of guidelines existence, only one-fifth reported satisfactory levels.[44,46] In terms of training received on safe medicines disposal from 13 studies, almost half (46.2%) reported it as insufficient.[26,27,29,33,43,46]

## Attitude domain

30 studies explored participants’ attitudes toward medication disposal.[16,18–42,49–52] The collective data from these studies fell into five sub-themes (see Figure 4). In 14 studies that examined attitudes toward protecting the environment and ensuring the safety of other living species, all reported a positive attitude.[16,18,19,22,26,27,31,32,34,36,39,41,49,50] Among the 16 studies that looked at giving appropriate advice on safe drug disposal, around two-thirds (n=11) had positive responses.[21,24,31,33,35,37,38,41,42,49,50] Of the eight studies focusing on providing services for drug disposal, 87.5% indicated a positive attitudes.[16,18,24,28,31,50,51] A strong inclination to engage in safe drug disposal practices by seeking further education or guidance was observed in 12 out of 13 studies (92.3%).[18,20,22,25,26,30,31,34,37,39,41,50] Finally, satisfaction with the current guidelines, legislation, or practices was mixed. Half of the four studies exhibited a positive response[50,51], while the remaining reflected dissatisfaction[24,36].

## Practice domain

35 studies evaluated participants’ behaviour in relation to medication disposal.[16,19–52] The studies examined different approaches to the disposal of medications, which were operationalised into two main practice domains and further into two subdomains (see Figure 5).

### Practice domain 1: Medicine disposal outside healthcare settings

Practice subdomain i): how healthcare professionals advised patients/clients on proper medication disposal

This subdomain comprised of 10 out of 35 studies (28.6%).[25,29,31,33–35,42,46,50,51] Results showed healthcare professionals provided a mixture of poor and appropriate advice to patients on medicines disposal.

Practice subdomain ii): how healthcare professionals disposed of their own unwanted medicines at home

The findings showed that most of the studies reflected inadequate practices, except for one[21]. Most of the participants were found to dispose of unwanted medications in household trash or flushed them down toilets.

### Practice domain 2: Medicine disposal within healthcare settings

The results revealed that healthcare professionals utilised different approaches to manage unwanted medicines in these settings, indicating a lack of awareness about proper disposal methods at their workplace. Some returned them to manufacturers, government agencies, or contractors, while others disposed of them through non-environmentally friendly means such as trash, sinks, or toilets.

### Studies assessed the practice of medicine disposal both within and outside healthcare settings

Two studies explored methods of disposal within and outside healthcare settings.[34,46] The results showed that patients were not receiving adequate or suitable guidance on how to dispose of medicines by healthcare staff. Similarly, staff employed various non-environmentally friendly methods within their workplace when disposing of medicines.

### Waste segregation into various dosage forms, controlled drugs, and outer packaging

Ten out of 35 studies (28.6%) examined the practice of disposal methods for different medication forms including solid, liquid, semi-solid, and parenteral.[17,19,21,22,25,30,34,37,38,52] Out of the ten studies, 90% indicated that the method of disposal was influenced by the dosage form of the medication.[17,19,21,22,25,34,37,38,52] Liquid forms were most disposed via sinks and solid forms were predominantly discarded in trash. Among four out of 35 studies (11.4%) investigated the disposal of controlled drugs.[21,34,37,52] In comparison to non-controlled drugs, all controlled drugs were reported safer disposal routes such as returning them to manufacturers, pharmacies, or government agencies instead of environmentally harmful options. Only three studies (8.6%) assessed participants on waste segregation practices such as utilising designated containers or separating outer packaging from the drugs.[37,48,49]

### Communication between patients and healthcare professionals

Only five out of 35 studies (14%) indicated healthcare staff had offered advice to patients on the proper disposal of medicines.[22,31,35,42,46] Interestingly, three studies (8.6%) mentioned healthcare professionals refusing to take back unwanted medicines from patients.[16,24,50] Four studies (11.4%) have indicated that patients seldom sought advice from healthcare professionals regarding the disposal of medicines.[28,31,33,46] A study conducted in Ireland revealed that patients were more inclined to return their medications to pharmacies when they received explicit instructions for proper disposal from healthcare professionals.[53] A study in India also supported healthcare professionals, encompassing doctors, nurses, and pharmacists seen by the public as crucial sources of information for guidance on medication disposal.[54]

## Legislations and take-back programs

The included studies highlighted the disparities in drug disposal legislation and guidelines across different countries (see Table 3 and Figure 6). Notably, seven countries (Bangladesh, Ethiopia, India, Iraq, Palestine, Saudi Arabia, Trinidad and Tobago) were found to lack any formal legislation or guidelines related to the disposal of unwanted medications.[17,19,20,26–28,34,36–39,41,48] These countries, representing various geographical regions and income groups, demonstrated a concerning gap in the regulation of drug disposal practices. In contrast, six countries (Brazil, Kosovo, Nigeria, Romania, South Africa, and the United Arab Emirates) have established legislation or guidelines governing drug disposal.[24,30,33,40,43,46,52] These guidelines, however, vary in their scope and enforcement, reflecting different national approaches to medication waste management. Interestingly, four countries (Indonesia, Kuwait, Malaysia, and Nepal) have instituted drug disposal guidelines that are applicable only within hospital settings, leaving community settings without clear regulations.[16,18,31,45,50] This highlights a significant gap, considering the widespread use of medications outside of healthcare facilities. The United States (US) is unique in having multiple laws and guidelines. A study in the US mentioned that opioids are considered suitable for flushing down the toilet by the Food and Drug Administration if proper disposal methods are not available.[35] In California specifically, the Department of Resources, Recycling, and Recovery advises against disposing any prescription or non-prescription substances down the drain or toilet.[35] It also suggests only throwing away medications in the trash when take-back or mail-back options are unavailable. These multiple regulations may cause confusion due to conflicting directives.[35] Moreover, five countries (Indonesia, Kosovo, Malaysia, Romania, and the United States) offer take-back programs for unused medications, an essential feature that provides an accessible means for safe disposal.[21,25,29,32,35,42,51] However, it is evident that these programs are not widely accessible globally.

## Obstacles to safe medication disposal

Out of 37 studies, 34 (92%) identified various obstacles to safe medication disposal (see Figure 7). [16,17,19–22,24–28,30–52] Twenty-eight studies (82%) highlighted issues related to insufficient knowledge, awareness, and training regarding proper medicine disposal methods.[17,20–22,24–27,30,32–36,38–41,43–52] Twenty-two studies (65%) emphasised the absence of services or established guidelines in this regard.[16,19,20,24,27,28,30–32,34–39,42,45,47,48,50–52] Even if there is a service in place such as a take-back program, almost one-third (10/34, 29%) highlighted limited accessibility to these services.[20,22,28,34,40,43,45,48,50,51] Six studies (18%) discussed concerns about lack of funding to sustain a service while relying on voluntary efforts.[24,28,30,31,50,51] A small number addressed fear of legal consequences (3%)[43], lack of interest from stakeholders (9%)[25,31,42], and insufficient time (6%)[35,42].

## Recommendations for improving medication disposal

All 37 studies have offered numerous recommendations for improving the proper disposal of medications, which were grouped into three levels: system, professionals, and patients (see Figure 8).

### System level

Twenty-five out of 37 studies (68%) advocate for government authorities to establish and enforce policies for medication disposal.[16,18–20,22–24,28,33,34,36,37,39–47,49–52] Twenty-one studies (57%) support the introduction of a drug take-back programme that allows patients to return their unused medications to local pharmacies for proper disposal.[16,18,19,22–24,26,30,31,34,36– 39,42,45,47,49–52] Twenty studies (54%) focus on increasing accessibility of take-back programme, gaining support from government initiatives, or securing funding to sustain the service.[17,20,22– 24,26–28,31,34,39–41,43,45,47,48,50–52]

### Professionals level

Twenty studies (54%) recommend expanding educational programmes on issues related to improper medication disposal and increasing awareness among all healthcare professionals about available guidelines and services for proper workplace disposal practices.[16,17,20,22,27–29,32– 36,38,40,45,46,48,49,51,52] Moreover, it is crucial to provide education on the harmful effects of medication waste on the environment before healthcare staff become qualified by integrating such knowledge into university curriculums (30%).[21,23,25,26,29,35,43,44,46,49,51] Two studies have mentioned collaborative work within multidisciplinary teams.[20,21] Four studies (11%) have recommended reducing overprescribing and limiting stock to prevent and minimise waste in the first instance.[23,24,30,39]

### Patients level

Seventeen studies (46%) recommend enhancing public education through awareness campaigns and utilising social media to promote proper medication disposal.[16,22–24,28,31,35,38,39,41–43,45– 47,51,52]

## Quality assessment

Majority of the studies were considered as good quality.[16–18,20–23,25,26,28,29,31,33– 36,38,39,41–43,45,49,50] It was noted that two of the studies performed less well with unclear descriptions of the study setting, uncertainty around the validity and reliability of the outcome measures employed, and a lack of appropriate statistical analysis.[24,44] See Table 4 and Figure 9 for the quality assessment of the individual studies.

## Discussion

### Knowledge of unwanted medicine disposal

The findings reveal significant areas of insufficient knowledge regarding the disposal of medications among healthcare professionals and students. Although there is a good level of awareness about the environmental impacts, there are significant gaps in knowledge regarding correct disposal methods, available services, guidelines, and training.[43] Improper disposal of medications remains a common problem among environmentally conscious individuals worldwide, indicating that environmental knowledge and awareness only partially influence medicine disposal behaviour.[55]

The lack of comprehensive knowledge and training among healthcare professionals is especially alarming because they have a crucial role in educating and influencing public behaviour on medication disposal. Due to insufficient guidance from healthcare professionals, the public often faces uncertainty and confusion regarding proper disposal methods.[56,57]

A study examined how participants perceived ecopharmacology and emphasised the importance of educating people about the emerging concept of ecopharmacovigilance.[26] However, previous studies have shown that environmental awareness did not have much influence on people’s drug disposal methods.[58,59]

The generally positive views on medication disposal from this review could be interpreted as, healthcare professionals and students’ preparedness to participate in, and improve disposal methods. However, some findings of reluctance of professional’s accountability and dissatisfaction with guidelines, show that additional measures are necessary to address these issues. Tai et al.[42] revealed pharmacists have a positive attitude and willingness to offer education on medication disposal. Some participants in the studies felt a sense of responsibility to provide guidance on medication disposal, as community healthcare professionals are often the initial point of contact for the public.[18,31,50] In contrast, other findings showed healthcare staff are hesitant to take on the full responsibility for advocating the return of unused medications.[23,27,36] This emphasises the importance of collaboration across multiple disciplines, involving patients, healthcare professionals, government bodies, healthcare institutions, and environmental agencies. Multiple collaboration with institutions is also supported in Abubakar et al.[60] in adverse drug reactions (ADR) detection, documentation, and reporting.

The results from the practice domain showed a notable difference in the practices pertinent to medicine disposal. Although there was a generally positive attitude and some understanding of appropriate disposal methods, the majority of participants did not follow the practice guidelines, especially outside healthcare settings. This inconsistency may be due to various factors such as inadequate practical training, limited resources, or unclear guidelines.

### Interrelationship between Knowledge of, Attitudes towards and Practices of medicine disposal

Knowledge influences attitudes as individuals who are informed about the hazards of improper disposal typically display a positive attitude towards environmentally friendly practices.[16,19,31,41] However, the extent and depth of this knowledge can vary significantly, which may account for the observed inconsistencies in attitudes and behaviours. The findings show that although there is a fair level of understanding regarding the environmental consequences of incorrect medication disposal, awareness does not consistently lead to appropriate disposal behaviours. For example, Srikanth et al.[41] showed that 93% participants agreed that unsafe disposal of unused medicines would adversely affect the environmental and human health. Nonetheless, 73% discard unwanted medicines in household trash, 7% dispose them in sinks, 6% flush them down toilets, and just 7% return them to pharmacies.[41]

According to Ajzen[61], positive attitudes towards a behaviour lead to better motivation and intention to engage in that behaviour. The results indicate a generally positive attitude towards the need for environmentally safe disposal practices. However, there is a gap between these positive attitudes and the actual practices, which could be due to several barriers including lack of confidence, fear of assuming responsibility, or perceived complexity of proper disposal methods.[23,24,27–29,36] These barriers indicate that although attitudes may align with knowledge, they do not necessarily overcome practical challenges that prevent these attitudes from translating into actions.[55] [62]

The reported medicines disposal practices in the findings, despite adequate environmental awareness and favourable attitudes, may be attributed to various factors such as lack of time, insufficient infrastructure for disposal, ambiguous guidelines, or limited focus on practical training in educational environments.[17,20,22,24,27,30–32,34–38,42–44,46,47,49–52] The findings underscore the necessity for a pragmatic approach that not only encourages knowledge and cultivates positive attitudes but also reinforces them with feasible, accessible, and user-friendly practices.[12]

Most of the studies in this review emphasised the widespread problem of insufficient understanding, awareness, and education about appropriate disposal methods.[16–52] This lack of knowledge and training indicates a larger systemic issue that impacts both healthcare professionals and the general population. Two previous studies have highlighted insufficient awareness of correct disposal methods and limited knowledge about drug take-back systems as factors contributing to low rates of medication return by the public.[63,64]

Additionally, almost two-thirds of the included studies indicated a lack of sufficient services or established protocols for medication disposal.[16,19,20,24,27,28,30–32,34–39,42,45,47,48,50–52] This insufficiency indicates a deficit in infrastructure and policy support needed to encourage appropriate disposal practices. The availability of programs like take-back initiatives was also found to be limited, with around one-third of the studies emphasising the restricted access to these services.[20,22,28,34,40,43,45,48,50,51]

Furthermore, there were references to legal concerns, limited interest among stakeholders, and time constraints.[25,31,42,43] While these issues are important to those impacted by them, they appear to be less widespread compared to challenges related to knowledge and access to services. This range of hurdles illustrates a healthcare system that recognises the importance of safe medication disposal but grapples with systemic barriers hindering the adoption of viable solutions.

The findings emphasise the need for a comprehensive approach involving education, infrastructure development, and policy implementation to promote environmental safety and public health. Ongoing professional development and targeted educational initiatives can help bridge the gap between knowledge and practice.[65] Improving general and specific knowledge can strengthen a positive attitude and in turn, lead to better practice.[12] Educational institutions should incorporate thorough training on medication disposal into the curricula for healthcare students, encompassing all aspects from environmental impacts to proper disposal methods and adherence to guidelines.[66]

Health authorities are encouraged to implement widespread public awareness initiatives to enhance overall understanding of proper medication disposal methods, with a focus on educating both healthcare professionals and the wider community. A previous study has shown providing legal backing for adverse drug reactions reporting and public awareness campaign are crucial to facilitate practice.[60]

Establishing accessible guidelines for medication disposal across various healthcare settings can standardise practices. The studies highlighted the importance of robust government policy implementation and enforcement, with two-thirds of the research supporting this approach to standardise medication disposal methods for environmental safety and public health.[16,18–20,22– 24,28,33,34,36,37,39–47,49–52]

Studies also emphasise the importance of ongoing government, environmental agencies assistance in enabling these efforts, demonstrating a comprehensive approach to addressing the challenges of disposing medications.[17,22,23,26–28,41,47,52] Healthcare professionals, regulatory agencies, and the pharmaceutical industry should collaborate to develop effective strategies for the proper disposal of medicines.[67] This approach not only focuses on the healthcare system but also involves community participation and policy interventions at a systemic level to create an efficient solution for this issue.[20,21]

## Geographical variation and economic impact

Our review encompassed studies from a variety of countries, revealing differences in healthcare infrastructure and levels of affluence that can impact the findings. High-income countries, such as the United States, generally have well-established medication disposal programs supported by government policies and initiatives, which facilitate safer disposal practices.[21,25,29,32,35,42,51] In contrast, middle and low-income countries like Brazil, Iraq, Bangladesh, and Ethiopia often encounter challenges in implementing safe medication disposal practices due to varying levels of infrastructure and support.[17,26,27,33,38] Furthermore, high, middle, and low-income countries alike have raised concerns that a lack of funding affects their ability to implement take-back programs.[26,27,30,38,46]

Awareness and education initiatives regarding proper medication disposal are present across high, middle, and low-income countries.[17,22,25,27–29,41,43,44,46,48] However, the prevalence and accessibility of educational programs and training vary. High-income countries typically have more systematic and widespread implementation of these programs, while low-income countries may struggle with infrastructure and resource challenges that affect the consistency and reach of educational efforts.[17,21,26,27,33,38] Addressing geographical and socioeconomic disparities is crucial for promoting better medication disposal practices globally among healthcare professionals and students. Tailored interventions and policies are necessary to enhance environmental safety and public health.

## Strength and limitations of the review

This review was designed using KAP model theoretical framework that helped researchers to look at evaluation of the studies more systematically. The KAP model helped structure our literature search and inclusion criteria, ensuring a comprehensive focus on studies addressing medication disposal. The framework provided a clear method for categorising and organising data, facilitating consistent data collection and analysis. During data synthesis, the KAP model enabled us to explore the relationships between KAP, offering deeper insights into the factors influencing medication disposal behaviours. In our discussion and recommendations, aligning our findings with the KAP framework allowed us to make targeted suggestions for improving practices, thus enhancing the credibility and relevance of our conclusions. This review included studies exploring the areas of knowledge, attitudes, and practices, without being limited to a combination of all three KAP model domains, allowing for the generalisability of the study findings and their potential impact on future research.

This review was restricted to studies published in English, which resulted in the exclusion of one study that was not in English. This limitation may have excluded relevant data published in other languages.

Two qualitative studies were included in this review, but their findings were presented as percentages and figures, which made it feasible for us to operationalise KAP.[50,51] For future research, if an in-depth analysis of qualitative studies is desired, appropriate qualitative methods should be employed.

The results only refer to two interventional studies, indicating that educational programs can positively impact proper medicine disposal.[29,48] There is limited knowledge about interventions that have been specifically implemented to address and improve medicine disposal practices.

## Future research

Authors encourage other researchers to adopt the current operationalisation knowledge, attitude, and practice domains to refine and enhance future studies. More work is needed to operationalise the practice components of disposing medicines such as exploring recycling and overprescribing. Future research should explore the importance of comprehensive educational approaches that target all aspects of KAP in order to bring about effective behavioural changes.

## Conclusion

This review investigated the knowledge, attitudes, and practices of healthcare staff and students regarding the discarding of unused medications. Overall, it was found that while healthcare staff and students have fair knowledge and positive attitudes toward medicine disposal, their actual practices are lacking. One significant challenge identified is the limited awareness about proper disposal methods coupled with a lack of established services or guidelines in many regions. Even in cases where take-back programs are available, they often face issues with accessibility. To tackle these challenges, it is suggested by the studies that governmental bodies should establish and enforce clear policies on medication disposal while also expanding educational initiatives to increase understanding among healthcare professionals and students about appropriate disposal methods. Furthermore, improving access to take-back programs is crucial for ensuring safe medication disposal and minimising potential environmental and health hazards.

## Supporting information

Appendix 1 PRISMA checklist

Protocol

Search strategies

Covidence extraction template

Abstraction table

## Data Availability

All data produced in the present work are contained in the manuscript

## Footnote

### Contributors

JL and KA conceived the idea and designed the protocol with support from the wider team. JL performed the literature search. DL, DN, RK and IMcG all supported JL with the design and conduct of the review. JL, DEA, ELL, and TA extracted the data and completed the quality assessment of the studies. Any queries were resolved through discussions with KA. JL and DEN drafted the first version of the manuscript with support from all other authors. KA reviewed the draft, with all other authors providing critical review. All authors read and approved the final manuscript.

### Funding

Whilst the review has received no direct funding, it has been undertaken as part of the Health Education England/ National Institute for Health Research Integrated Clinical Academic (ICA) programme award. KA is in part supported by the National Institute for Health and Care Research (NIHR) Applied Research Collaboration (ARC) Northwest London. The views expressed are those of the author and not necessarily those of the NIHR or the Department of Health and Social Care. Funding to pay the Open Access publication charges for this article was provided by the Imperial Open Access Fund (Grant Number: N/A).

### Patient and public involvement

Patients and/or the public were not involved in the design, or conduct, or reporting, or dissemination plans of this research.

### Provenance and peer review

Not commissioned; externally peer reviewed.

### Data sharing statement

No additional data are available.

