## Supplementary material for "Assessing the knowledge, attitudes and practices of healthcare staff and students regarding disposal of unwanted medications: A systematic review": Protocol

Review methods were amended after registration. Please see the revision notes and previous versions for detail.

### Citation

Janeme Lam, Dayana El Nsouli, E Lyn Lee, Tawfiq Alqeisi, Keivan Ahmadi. Assessing the knowledge, attitudes and practices of healthcare staff and students regarding disposal of unwanted medications- A systematic review. PROSPERO 2024 CRD42024503162 Available from:
https://www.crd.york.ac.uk/prospero/display_record.php?ID=CRD42024503162

### Review question

What is the level of knowledge, attitudes, practices and perceptions of the healthcare staff and the healthcare students about disposal of unwanted medications?

-P: Pharmacists, doctors, nurses, medical students, pharmacy students, nursing students

-I: Knowledge and/or attitudes and/or perceptions, and/or practices about medicines waste disposal

-C: Not applicable

-O: Knowledge, attitudes, practices, and/or perceptions of medicines waste disposal

### Searches

The following databases will be searched: MEDLINE, Embase, PsycINFO, CINAHL, Web of Science and Google Scholar. Manual searches of reference lists of included articles will be conducted to identify further papers. Systematic reviews will be excluded but their included studies and reference lists will be searched for additional pertinent literature.

- Publication dates: no restriction

Restrictions:

- Language: English language only

### Types of study to be included

- Inclusion: The review will include qualitative, quantitative, and mixed-method primary research studies.

- Exclusion: Any type of reviews including scoping reviews, narrative reviews, umbrella reviews, systematic reviews, or meta-analysis studies. Any non-peer reviewed publications such as reports, audits, and podcasts.

### Condition or domain being studied

Leftover or unwanted medicines include all medications no longer being used for the initial prescribed indication, accounting for dominating cause of environmental contamination because of improper disposal. Healthcare professionals need to monitor the effects of drugs not only as a good medical practice, but also to safeguard the environment. The presence of pharmaceuticals in the environment poses a potential direct, and indirect risk to humans and the ecosystem. These pharmaceuticals enter the environment through various routes such as drugs being excreted after consumption or when unused medicines are discarded improperly. If the environmental damage is not being addressed, it will have a huge impact in causing species to go extinct in an accelerated rate and disruption of the food chain. Healthcare professionals and future healthcare professionals should be aware of the safe disposal of medicines and ecopharmacovigilance which can be defined as science and activities concerning detection, assessment, understanding and prevention of adverse effects related to the presence of pharmaceuticals in the environment.

### Participants/population

- Inclusion: Pharmacists, doctors, nurses, medical students, pharmacy students, nursing students.

- Exclusion: Other healthcare professions such as midwives, occupational therapists, physiotherapists, radiotherapists, dentists, or allied healthcare professions that do not have significant input dealing with medicines disposal. Students who are not medical, pharmacy, or nursing background.

### Intervention(s), exposure(s)

- Inclusion: Knowledge, attitudes, perceptions, and/or practices about medicines waste disposal.

- Exclusion: Studies not investigating knowledge, attitudes, perceptions, and/or practices about medicines waste disposal.

### Comparator(s)/control

Not applicable

### Context

Studies will include any primary, secondary, or tertiary settings. Pharmacy, medical or nursing students, including any stage of undergraduate or postgraduate training.

### Main outcome(s)

The primary outcome is to measure the levels of the healthcare staff and students' knowledge, as well as their attitudes, and practices about disposal of unwanted medications.

### Additional outcome(s)

Additional outcome of the review will be to investigate any validated measures to assess healthcare staff and students' knowledge, attitudes, and practices about medicines disposal in the existing literature.

### Data extraction (selection and coding) [1 change]

Two team members (JL and DEN) will screen independently titles and abstracts for inclusion. Full text articles will then be screened and assessed for inclusion. Any disagreement between reviewers over the eligibility of particular studies will be resolved through discussion with a third reviewer (KA).

Covidence software will be used as platform to carry out primary screening and data extractions. A data extraction form will be adapted from Cochrane's Data Extraction Template. Data extraction will be carried out by four of the researchers (JL, DEN, ELL, TA) independently. Any discrepancies for the data extraction amongst the researchers will be resolved by KA.

Information to be extracted include:

Study title, the first author, study duration, study participants (type of professions or students), sample size of each profession, participants level of qualification (healthcare staff or healthcare students), study setting, study design, domains of knowledge/attitudes/practices of medicines disposal, findings, reported limitations, and conclusions.

### Risk of bias (quality) assessment [1 change]

The methodological quality of each study will be assessed using an appropriate Joanna Briggs Institute's critical appraisal tool. Methodological quality for each study is reported as high, medium, or low using the overall score generated. The level of quality will not be a reason for exclusion of the studies. Four team members (JL, DEN, ELL, TA) will independently assess the quality of the studies. Any discrepancies for the data extraction amongst the researchers will be resolved by a fifth reviewer KA. Each question will require a yes, no, or unclear answer. Studies receive 1 point for each "yes" response and 0 points for "no" or "unclear." Based on the total score of 5, studies are categorised as 'good quality' if scoring between 4-5, 'fair' if scoring a total of 3, and 'low' if the score is between 0-2.

### Strategy for data synthesis

Quantitative data synthesis:

A narrative or descriptive synthesis will be sought regarding the knowledge, attitudes, and practices of healthcare staff and students regarding medicines disposal. It is anticipated that we could adapt a numerical system , most probably in percentage, by referring to the valid and reliable tools such as the questionnaires developed by Amod F, Chetty K, Essa AS, Hlela L, Maharaj C, Oosthuizen F. A pilot study to determine public trends in storage and disposal of medicines. SAPJ. 2008;75:7; and Seehusen DA, Edwards J. Patient practices and beliefs concerning disposal of medications. J Am Board Fam Med. 2006;19:542–7 to measure levels of knowledge regarding the medicine disposal. Such data will be tabulated in three main domains i.e., Knowledge, Attitudes, and Practices for further analysis. The outcomes will be reported using descriptive as well as inferential analysis if the assumptions are hold.

Qualitative data synthesis:

Qualitative data will be summarised according to the objectives of this study by using a narrative description of the available evidence, gathered from qualitative studies' findings such as quotes, themes reported from the quotes, etc. Themes and quotes will be categorised based on the above-mentioned three main domains i.e., Knowledge, Attitudes and Practices to capture rich data pertinent to each domain

Integration of the Quantitative data and the Qualitative data:

The analyses will be integrated to create a full picture on the reviewed evidence (Quantitative and Qualitative) regarding the healthcare professionals' and healthcare students' of knowledge, attitudes, and practices regarding the medicines disposal.

### Analysis of subgroups or subsets

If sufficient data is available, a subgroup analysis will be conducted for the main outcome by countries, professional backgrounds, difference between students and professionals and study settings. Any subgroups or subsets will be identified and reported descriptively.

### Contact details for further information

Janeme Lam

### Organisational affiliation of the review

University of Lincoln

https://www.lincoln.ac.uk/home/

### Review team members and their organisational affiliations [1 change]

Miss Janeme Lam. Pharmacy Department, Northampton General Hospital, Northampton, UK

Miss Dayana El Nsouli. University Hospitals of Derby and Burton, Derby, UK

E Lyn Lee. School of Pharmacy, IMU University, 57000, Kuala Lumpur, Malaysia

Mr Tawfiq Alqeisi. Queens Medical Centre, Nottingham University Hospitals, Nottingham, UK

Dr Keivan Ahmadi. Imperial College London, London, UK

### Collaborators

Dr Ian McGonagle. School of Health and Social Care, University of Lincoln, Lincoln, UK

Dr Ros Kane. School of Health and Social Care, University of Lincoln, Lincoln, UK

Dr David Nelson. Lincoln International Institute for Rural Health (LIIRH), University of Lincoln, Lincoln, UK

Ms Despina Laparidou. School of Health and Social Care, University of Lincoln, Lincoln, UK

Dr Samuel Cooke. Lincoln International Institute for Rural Health (LIIRH), University of Lincoln, Lincoln, UK

### Type and method of review

Narrative synthesis, Systematic review

### Anticipated or actual start date

01 February 2024

### Anticipated completion date

01 February 2025

### Funding sources/sponsors

Whilst the review has received no direct funding, it has been undertaken as part of the HEE/NIHR Integrated Clinical Academic (ICA) programme award.

### Conflicts of interest

### Language

English

### Country

England

### Stage of review

Review Ongoing

### Subject index terms status

Subject indexing assigned by CRD

### Subject index terms

MeSH headings have not been applied to this record

### Date of registration in PROSPERO

01 February 2024

### Date of first submission

01 February 2024

### Stage of review at time of this submission [3 changes]

| Stage | Started | Completed |
|---|---|---|
| Preliminary searches | Yes | Yes |
| Piloting of the study selection process | Yes | Yes |
| Formal screening of search results against eligibility criteria | Yes | Yes |
| Data extraction | Yes | Yes |
| Risk of bias (quality) assessment | Yes | Yes |
| Data analysis | Yes | Yes |

### Revision note

We have changed the title slightly to better reflect the content of our study.

*The record owner confirms that the information they have supplied for this submission is accurate and complete and they understand that deliberate provision of inaccurate information or omission of data may be construed as scientific misconduct.*

*The record owner confirms that they will update the status of the review when it is completed and will add publication details in due course.*

### Versions

01 February 2024

01 February 2024

05 June 2024

18 November 2024
