## Supplementary material for "Assessing the knowledge, attitudes and practices of healthcare staff and students regarding disposal of unwanted medications: A systematic review": Search strategies

**Appendix 3**

**Search Strategies**

Database searched:

| **Database** | **No. of Hits** | **Date last searched** |
| --- | --- | --- |
| Medline | 145 | 1/2/2024 |
| Embase | 358 | 1/2/2024 |
| Web of Science | 200 | 31/1/2024 |
| Cinahl | 94 | 31/1/2024 |
| PsycINFO | 18 | 31/1/2024 |
| Google Scholar | 221 | 23/2/2024 |

Search Strategy for each database:

**Database: Ovid Medline** <no restriction on date>

Last searched: 1/2/2024

| **Set** | **Search statement** | **Results** |
| --- | --- | --- |
| 1 | "pharmacist*".ab,ti. | 44218 |
| 2 | exp Pharmacists/ | 22314 |
| 3 | "pharmacy student*".ab,ti. | 3887 |
| 4 | "student* pharmacist*".ab,ti. | 945 |
| 5 | exp Students, Pharmacy/ | 4599 |
| 6 | "doctor*".ab,ti. | 155469 |
| 7 | exp Physicians/ | 182620 |
| 8 | "medical student*".ab,ti. | 54509 |
| 9 | exp Students, Medical/ | 44710 |
| 10 | "nurse*".ab,ti. | 328223 |
| 11 | exp Nurses/ | 99909 |
| 12 | "nursing student*".ab,ti. | 21535 |
| 13 | "student* nurse*".ab,ti. | 4740 |
| 14 | exp Students, Nursing/ | 31228 |
| 15 | 1 or 2 or 3 or 4 or 5 or 6 or 7 or 8 or 9 or 10 or 11 or 12 or 13 or 14 | 780343 |
| 16 | (medicine* adj3 disposal*).ab,ti. | 91 |
| 17 | (pharmaceutic* adj3 disposal*).ab,ti. | 109 |
| 18 | (medication* adj3 disposal*).ab,ti. | 230 |
| 19 | (Medicine* adj3 dispose*).ab,ti. | 31 |
| 20 | (pharmaceutic* adj3 dispose*).ab,ti. | 14 |
| 21 | (Medication* adj3 dispose*).ab,ti. | 86 |
| 22 | (unwanted adj3 medicine*).ab,ti. | 43 |
| 23 | (unwanted adj3 medication*).ab,ti. | 82 |
| 24 | (unwanted adj3 pharmaceutic*).ab,ti. | 11 |
| 25 | (expire* adj3 medicine*).ab,ti. | 108 |
| 26 | (expire* adj3 medication*).ab,ti. | 132 |
| 27 | (expire* adj3 pharmaceutic*).ab,ti. | 41 |
| 28 | (drug* adj3 disposal*).ab,ti. | 253 |
| 29 | (drug* adj3 dispose*).ab,ti. | 52 |
| 30 | (unwanted adj3 drug*).ab,ti. | 429 |
| 31 | (expire* adj3 drug*).ab,ti. | 184 |
| 32 | 16 or 17 or 18 or 19 or 20 or 21 or 22 or 23 or 24 or 25 or 26 or 27 or 28 or 29 or 30 or 31 | 1463 |
| 33 | "knowledge*".ab,ti. | 958705 |
| 34 | exp Knowledge/ | 14951 |
| 35 | "attitude*".ab,ti. | 196166 |
| 36 | "practice*".ab,ti. | 1144638 |
| 37 | "practise*".ab,ti. | 11886 |
| 38 | "perception*".ab,ti. | 338697 |
| 39 | exp Attitude/ | 642270 |
| 40 | exp Perception/ | 485012 |
| 41 | 33 or 34 or 35 or 36 or 37 or 38 or 39 or 40 | 3093356 |
| 42 | 15 and 32 and 41 | **145** |

**Database: Ovid Embase** <no restriction on date>

Last searched: 1/2/2024

| **Set** | **Search statement** | **Results** |
| --- | --- | --- |
| 1 | "pharmacist*".ab,ti. | 94235 |
| 2 | exp community pharmacist/ or exp clinical pharmacist/ or exp pharmacist/ or exp hospital pharmacist/ | 101038 |
| 3 | "pharmacy student*".ab,ti. | 6912 |
| 4 | "student* pharmacist*".ab,ti. | 1778 |
| 5 | exp pharmacy student/ | 9812 |
| 6 | "doctor*".ab,ti. | 223362 |
| 7 | exp general practitioner/ or exp physician/ | 995030 |
| 8 | "medical student*".ab,ti. | 72730 |
| 9 | exp medical student/ | 92769 |
| 10 | "nurse*".ab,ti. | 396146 |
| 11 | exp registered nurse/ or exp nurse/ or exp staff nurse/ | 222593 |
| 12 | "nursing student*".ab,ti. | 21545 |
| 13 | "student* nurse*".ab,ti. | 4435 |
| 14 | exp nursing student/ | 33216 |
| 15 | 1 or 2 or 3 or 4 or 5 or 6 or 7 or 8 or 9 or 10 or 11 or 12 or 13 or 14 | 1716465 |
| 16 | (medicine* adj3 disposal*).ab,ti. | 175 |
| 17 | (pharmaceutic* adj3 disposal*).ab,ti. | 147 |
| 18 | (medication* adj3 disposal*).ab,ti. | 398 |
| 19 | (Medicine* adj3 dispose*).ab,ti. | 62 |
| 20 | (pharmaceutic* adj3 dispose*).ab,ti. | 19 |
| 21 | (Medication* adj3 dispose*).ab,ti. | 142 |
| 22 | (unwanted adj3 medicine*).ab,ti. | 73 |
| 23 | (unwanted adj3 medication*).ab,ti. | 132 |
| 24 | (unwanted adj3 pharmaceutic*).ab,ti. | 27 |
| 25 | (expire* adj3 medicine*).ab,ti. | 200 |
| 26 | (expire* adj3 medication*).ab,ti. | 241 |
| 27 | (expire* adj3 pharmaceutic*).ab,ti. | 60 |
| 28 | (drug* adj3 disposal*).ab,ti. | 408 |
| 29 | (drug* adj3 dispose*).ab,ti. | 87 |
| 30 | (unwanted adj3 drug*).ab,ti. | 609 |
| 31 | (expire* adj3 drug*).ab,ti. | 352 |
| 32 | 16 or 17 or 18 or 19 or 20 or 21 or 22 or 23 or 24 or 25 or 26 or 27 or 28 or 29 or 30 or 31 | 2392 |
| 33 | "knowledge*".ab,ti. | 1196430 |
| 34 | exp knowledge/ | 214077 |
| 35 | "attitude*".ab,ti. | 242077 |
| 36 | "practice*".ab,ti. | 1526899 |
| 37 | "practise*".ab,ti. | 18244 |
| 38 | "perception*".ab,ti. | 410097 |
| 39 | exp attitude/ | 956724 |
| 40 | exp perception/ | 522559 |
| 41 | 33 or 34 or 35 or 36 or 37 or 38 or 39 or 40 | 3969650 |
| 42 | 15 and 32 and 41 | **358** |

**Database: Clarivate Web of Science** <no restriction on date>

Last searched: 31/1/2024

| **Set** | **Search statement** | **Results** |
| --- | --- | --- |
| 1 | TS=(pharmacist*) | 52259 |
| 2 | TS=(pharmacy student*) | 10521 |
| 3 | TS=(student* pharmacist*) | 4186 |
| 4 | TS=(doctor*) | 176151 |
| 5 | TS=(physician*) | 416539 |
| 6 | TS=(medical student*) | 101459 |
| 7 | TS=(nurse*) | 274620 |
| 8 | TS=(nursing student*) | 42845 |
| 9 | TS=(student* nurse*) | 24460 |
| 10 | #9 OR #8 OR #7 OR #6 OR #5 OR #4 OR #3 OR #2 OR #1 | 927716 |
| 11 | TS=(medicine* near/3 disposal*) | 174 |
| 12 | TS=(pharmaceutic* near/3 disposal*) | 213 |
| 13 | TS=(medication* near/3 disposal*) | 293 |
| 14 | TS=(medicine* near/3 dispose*) | 50 |
| 15 | TS=(pharmaceutic* near/3 dispose*) | 34 |
| 16 | TS=(medication* near/3 dispose*) | 105 |
| 17 | TS=(unwanted near/3 medicine*) | 71 |
| 18 | TS=(unwanted near/3 medication*) | 128 |
| 19 | TS=(unwanted near/3 pharmaceutic*) | 30 |
| 20 | TS=(expire* near/3 medicine*) | 183 |
| 21 | TS=(expire* near/3 medication*) | 168 |
| 22 | TS=(expire* near/3 pharmaceutic*) | 74 |
| 23 | TS=(drug* near/3 disposal*) | 367 |
| 24 | TS=(drug* near/3 dispose*) | 60 |
| 25 | TS=(unwanted near/3 drug*) | 573 |
| 26 | TS=(expire* near/3 drug*) | 348 |
| 27 | #26 OR #25 OR #24 OR #23 OR #22 OR #21 OR #20 OR #19 OR #18 OR #17 OR #16 OR #15 OR #14 OR #13 OR #12 OR #11 | 2188 |
| 28 | TS=(knowledge*) | 2037086 |
| 29 | TS=(attitude*) | 535226 |
| 30 | TS=(practice*) | 2080594 |
| 31 | TS=(practise*) | 20791 |
| 32 | TS=(perception*) | 871985 |
| 33 | #32 OR #31 OR #30 OR #29 OR #28 | 4840560 |
| 34 | #33 AND #27 AND #10 | **200** |

**Database: EBSCOhost CINAHL** <no restriction on date>

Last searched: 31/1/2024

| **Set** | **Search statement** | **Results** |
| --- | --- | --- |
| 1 | TI pharmacist* OR AB pharmacist* | 23419 |
| 2 | (MH "Pharmacists") | 18508 |
| 3 | "pharmacy student*" | 1498 |
| 4 | TI student* pharmacist* OR AB student* pharmacist* | 283 |
| 5 | (MH "Students, Pharmacy") | 1627 |
| 6 | TI doctor* OR AB doctor* | 71660 |
| 7 | (MH "Physicians+") | 133082 |
| 8 | TI medical student* OR AB medical student* | 18607 |
| 9 | (MH "Students, Medical+") | 30039 |
| 10 | TI nurse* OR AB nurse* | 384345 |
| 11 | (MH "Nurses+") | 230591 |
| 12 | TI nursing student* OR AB nursing student* | 27819 |
| 13 | TI student* nurse* OR AB student* nurse* | 5500 |
| 14 | (MH "Students, Nursing+") | 45530 |
| 15 | S1 OR S2 OR S3 OR S4 OR S5 OR S6 OR S7 OR S8 OR S9 OR S10 OR S11 OR S12 OR S13 OR S14 | 737129 |
| 16 | TI medicine* n3 disposal* OR AB medicine* n3 disposal* | 49 |
| 17 | TI pharmaceutic* n3 disposal* OR AB pharmaceutic* n3 disposal* | 43 |
| 18 | TI medication* n3 disposal* OR AB medication* n3 disposal* | 181 |
| 19 | TI medicine* n3 dispose* OR AB medicine* n3 dispose* | 17 |
| 20 | TI pharmaceutic* n3 dispose* OR AB pharmaceutic* n3 dispose* | 8 |
| 21 | TI medication* n3 dispose* OR AB medication* n3 dispose* | 55 |
| 22 | TI unwanted n3 medicine* OR AB unwanted n3 medicine* | 16 |
| 23 | TI unwanted n3 medication* OR AB unwanted n3 medication* | 63 |
| 24 | TI unwanted n3 pharmaceutic* OR AB unwanted n3 pharmaceutic* | 4 |
| 25 | TI expire* n3 medicine* OR AB expire* n3 medicine* | 42 |
| 26 | TI expire* n3 medication* OR AB expire* n3 medication* | 74 |
| 27 | TX expire* n3 pharmaceutic* OR AB expire* n3 pharmaceutic* | 15 |
| 28 | TX drug* n3 disposal* OR AB drug* n3 disposal* | 165 |
| 29 | TX drug* n3 dispose* OR AB drug* n3 dispose* | 37 |
| 30 | TX unwanted n3 drug* OR AB unwanted n3 drug* | 103 |
| 31 | TX expire* n3 drug* OR AB expire*n3 drug* | 66 |
| 32 | S16 OR S17 OR S18 OR S19 OR S20 OR S21 OR S22 OR S23 OR S24 OR S25 OR S26 OR S27 OR S28 OR S29 OR S30 OR S31 | 734 |
| 33 | TI knowledge* OR AB knowledge* | 281507 |
| 34 | (MH "Knowledge+") | 91037 |
| 35 | TI attitude* OR AB attitude* | 100311 |
| 36 | TI practice* OR AB practice* | 581764 |
| 37 | TI practise* OR AB practise* | 5498 |
| 38 | TI perception* OR AB perception* | 156421 |
| 39 | (MH "Attitude+") | 548965 |
| 40 | (MH "Perception+") | 93801 |
| 41 | S33 OR S34 OR S35 OR S36 OR S37 OR S38 OR S39 OR S40 | 1364617 |
| 42 | S15 AND S32 AND S41 | **94** |

**Database: ProQuest PsycINFO** <no restriction on date>

Last searched: 31/1/2024

| **Set** | **Search statement** | **Results** |
| --- | --- | --- |
| 1 | tiab(pharmacist*) | 3889 |
| 2 | MAINSUBJECT.EXACT.EXPLODE("Pharmacists") | 1990 |
| 3 | tiab("pharmacy student*") | 430 |
| 4 | tiab("student* pharmacist*") | 51 |
| 5 | tiab(doctor*) | 40966 |
| 6 | MAINSUBJECT.EXACT.EXPLODE("Physicians") | 49952 |
| 7 | tiab("medical student*") | 14377 |
| 8 | MAINSUBJECT.EXACT.EXPLODE("Medical Students") | 15288 |
| 9 | tiab(nurse*) | 75677 |
| 10 | MAINSUBJECT.EXACT.EXPLODE("Nurses") | 39489 |
| 11 | tiab("nursing student*") | 6763 |
| 12 | tiab("student* nurse*") | 1616 |
| 13 | MAINSUBJECT.EXACT.EXPLODE("Nursing Students") | 6976 |
| 14 | [S1] OR [S2] OR [S3] OR [S4] OR [S5] OR [S6] OR [S7] OR [S8] OR [S9] OR [S10] OR [S11] OR [S12] OR [S13] | 177710 |
| 15 | tiab(medicine* near/3 disposal*) | 6 |
| 16 | tiab(pharmaceutic* NEAR/3 disposal*) | 1 |
| 17 | tiab(medication* NEAR/3 disposal*) | 44 |
| 18 | tiab(medicine* NEAR/3 dispose*) | 2 |
| 19 | tiab(pharmaceutic* NEAR/3 dispose*) | 2 |
| 20 | tiab(medication* NEAR/3 dispose*) | 14 |
| 21 | tiab(unwanted NEAR/3 medicine*) | 4 |
| 22 | tiab(unwanted NEAR/3 medication*) | 38 |
| 23 | tiab(unwanted NEAR/3 pharmaceutic*) | 1 |
| 24 | tiab(expire* NEAR/3 medicine*) | 3 |
| 25 | tiab(expire* NEAR/3 medication*) | 14 |
| 26 | tiab(expire* NEAR/3 pharmaceutic*) | 1 |
| 27 | tiab(drug* NEAR/3 disposal*) | 37 |
| 28 | tiab(drug* NEAR/3 dispose*) | 17 |
| 29 | tiab(unwanted* NEAR/3 drug*) | 64 |
| 30 | tiab(expire* NEAR/3 drug*) | 21 |
| 31 | [S15] OR [S16] OR [S17] OR [S18] OR [S19] OR [S20] OR [S21] OR [S22] OR [S23] OR [S24] OR [S25] OR [S26] OR [S27] OR [S28] OR [S29] OR [S30] | 220 |
| 32 | tiab(knowledge*) | 323629 |
| 33 | MAINSUBJECT.EXACT.EXPLODE("Knowledge (General)") | 1221824 |
| 34 | tiab(attitude*) | 218613 |
| 35 | MAINSUBJECT.EXACT.EXPLODE("Attitudes") | 504981 |
| 36 | tiab(practice*) | 493111 |
| 37 | tiab(practise*) | 3156 |
| 38 | MAINSUBJECT.EXACT.EXPLODE("Practice") | 11247 |
| 39 | tiab(perception*) | 332527 |
| 40 | MAINSUBJECT.EXACT.EXPLODE("Social Perception") | 88794 |
| 41 | [S32] OR [S33] OR [S34] OR [S35] OR [S36] OR [S37] OR [S38] OR [S39] OR [S40] | 1520903 |
| 42 | [S14] AND [S31] AND [S41] | **18** |

**Database: Google Scholar** <no restriction on date>

Last searched: 23/2/2024

medicine disposal knowledge attitude practice pharmacist doctor nurse student	**221**

The first 200 articles (first 20 pages) were retrieved.
