## Supplementary material for "Assessing the knowledge, attitudes and practices of healthcare staff and students regarding disposal of unwanted medications: A systematic review": Covidence extraction template

**Appendix 4**

**Covidence™ extraction template**

|  | **Reviewer 1** | **Reviewer 2** |
| --- | --- | --- |
| **Study ID** |  |  |
| **Title** |  |  |
| **Lead author name** |  |  |
| **Year of study** |  |  |
| **Country in which the study conducted** |  |  |
| **Legislation or guidelines on medicines disposal in the geographical location** |  |  |
| **Aim and objectives of study** |  |  |
| **Study design** |  |  |
| **Data collection tool** |  |  |
| **When study was conducted** |  |  |
| **duration of the study** |  |  |
| **Study setting** |  |  |
| **Total number of participants** |  |  |
| **Population description (healthcare staff and/or students assessed)** |  |  |
| **Profession of staff/students and number of participants in each subgroup** |  |  |
| **Mean age or range of participants** |  |  |
| **Gender of participants** |  |  |
| **Inclusion criteria** |  |  |
| **Exclusion criteria** |  |  |
| **Main findings of knowledge** |  |  |
| **Main findings of attitude** |  |  |
| **Main findings of practice** |  |  |
| **Patient education** |  |  |
| **Environmental element** |  |  |
| **Current challenges reported by respondents or article (barriers)** |  |  |
| **Other comments or future recommendations by respondents or article (enablers)** |  |  |
