## Supplementary material for "Assessing the knowledge, attitudes and practices of healthcare staff and students regarding disposal of unwanted medications: A systematic review": Figures and tables

**Data supplement 5 - Online figures**

### Figure 1 PRISMA diagram of the systematic review

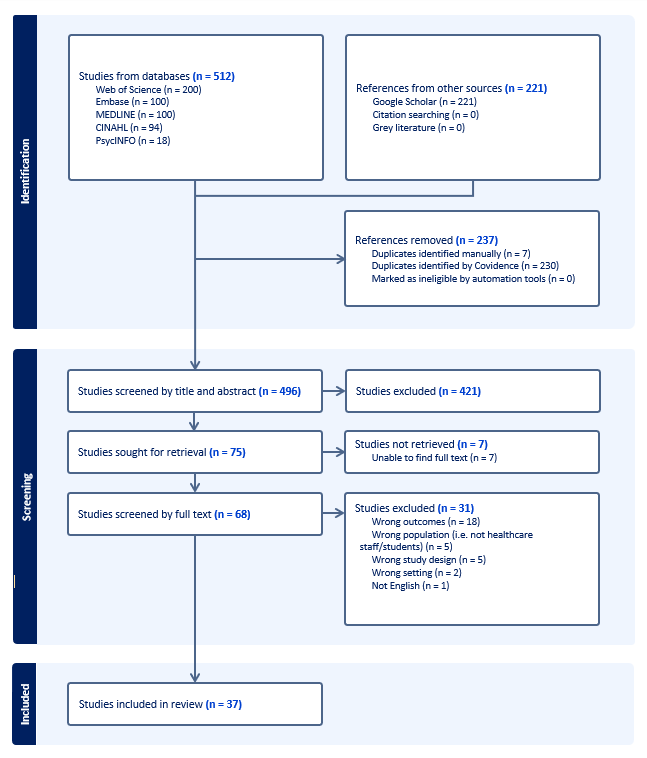

### Figure 2 Country of origin of the articles included in this review

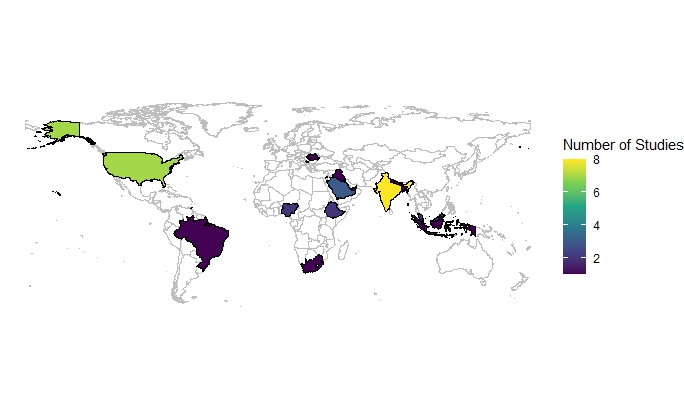

### Figure 3 Levels of awareness and training regarding medicine disposal among participants as reported in studies

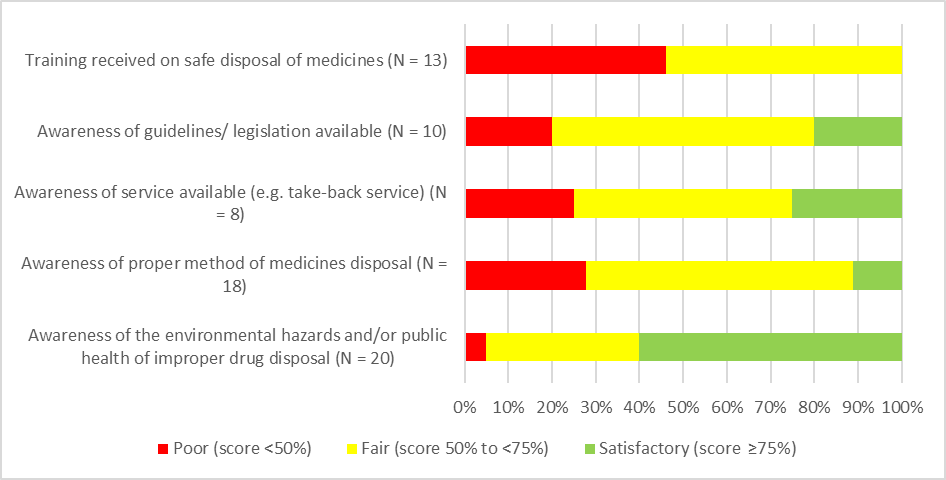

### Figure 4 Levels of attitude regarding medicine disposal among participants as reported in studies

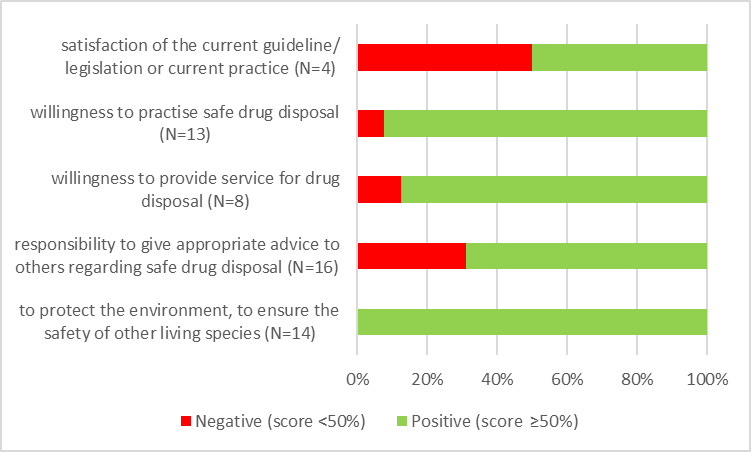

### Figure 5 Operationalisation of Practice domain

Practice- methods of disposal

Outside healthcare settings

Advice from HCPs

HCPs' personal disposal practice

Within healthcare settings

Abbreviation: HCPs= healthcare professionals

### Figure 6 Legislation/ guideline availability regarding medicine disposal as reported in studies

No. of studies

|  | no. of studies |
| --- | --- |
| No legislation/ guideline available | 7 |
| Legislation/ guideline available | 6 |
| Legislation/ guideline available in hospital but not in community setting | 4 |
| Multiple legislations/ guidelines available | 1 |

### Figure 7 Challenges regarding medicine disposal among participants as reported in studies

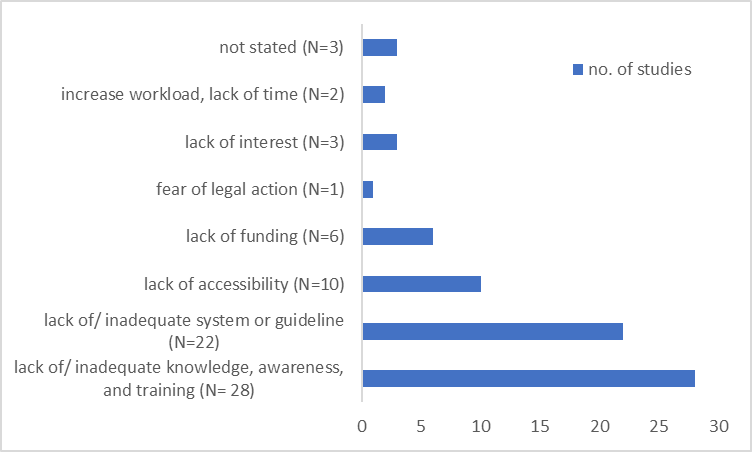

### Figure 8 Recommendations regarding medicine disposal among participants as reported in studies

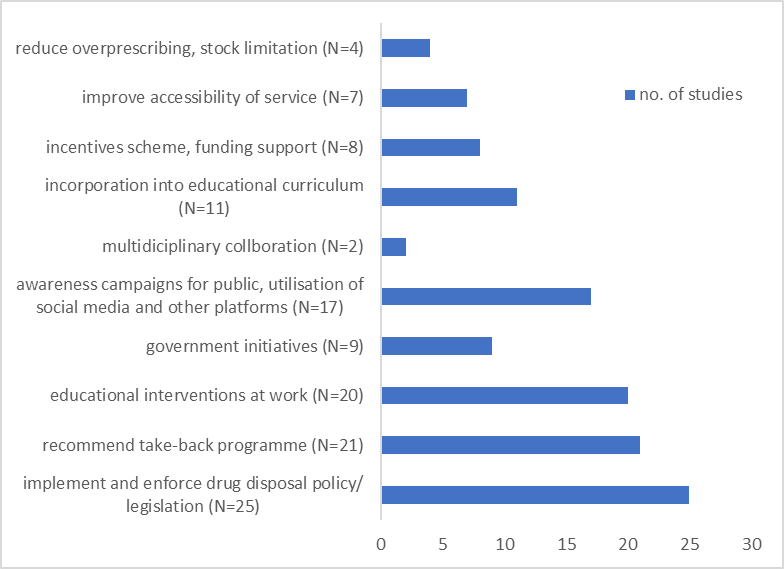

### Figure 9 Quality assessment of the included studies

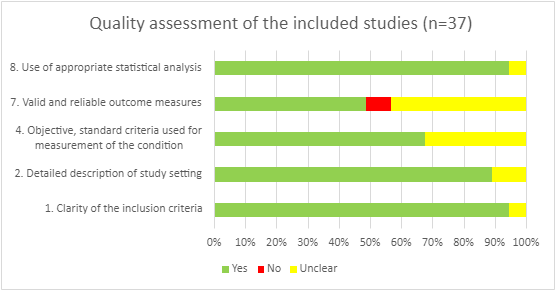

### Table 1 Inclusion and Exclusion criteria

| Criterion | Inclusion | Exclusion |
| --- | --- | --- |
| Type of studies | Qualitative, quantitative, and mixed-method primary research studies | Any type of reviews including scoping reviews, narrative reviews, umbrella reviews, systematic reviews, meta-analysis studies;  any non-peer reviewed publications such as reports, audits, and podcasts. |
| Types of participants | Pharmacists, doctors, nurses, medical students, pharmacy students, nursing students including any stage of undergraduate or postgraduate training | Patients or other healthcare professions such as midwives, occupational therapists, physiotherapists, radiotherapists, dentists, or allied healthcare professions that do not have significant input dealing with medicines disposal;  students who are not medical, pharmacy, or nursing background. |
| Time period | Any | - |
| Geographical location | Any | - |
| Setting | Primary, secondary, or tertiary care | - |
| Language | English | Other languages than English |
| Study focus | Studies that focus on knowledge, attitudes, and/or practices of healthcare staff or students about medicines waste disposal. | Studies not investigating knowledge, attitudes, and/or practices about medicines waste disposal. |

### Table 2 Overview of databases searched

| Name of database | Contents | Platform/interface |
| --- | --- | --- |
| MEDLINE | Journals related to life sciences, particularly biomedicine. | Ovid |
| Embase | Journals related to life sciences, particularly biomedicine. | Ovid |
| Cumulative Index to Nursing and Allied Health Literature (CINAHL) | Journals related to nursing and allied health issues. | EBSCOhost |
| Web of Science | A multi-disciplinary database containing journals related to medical and social issues among others. | Clarivate |
| PsycINFO | Peer-reviewed literature related to mental health and the behavioural sciences. | ProQuest |
| Google Scholar | Academic literature across a range of publishing formats and disciplines. | <https://scholar.google.com/> |

### Table 3 Legislation/ guideline availability regarding medicine disposal as reported in studies

| **No legislations/ guidelines available (n=7)** | **Legislations/ guidelines available (n=6)** | **Legislations/ guidelines available in hospitals but not in community settings (n=4)** | **Multiple legislations/ guidelines available (n=1)** | **Take-back program available (n=5)** |
| --- | --- | --- | --- | --- |
| Bangladesh ^b^ | Brazil ^c^ | Indonesia ^c^ | United States ^d^ | Indonesia |
| Ethiopia ^a^ | Kosovo ^c^ | Kuwait ^d^ |  | Kosovo |
| India ^b^ | Nigeria ^b^ | Malaysia ^c^ |  | Malaysia |
| Iraq ^c^ | Romania ^d^ | Nepal ^b^ |  | Romania |
| Palestine ^c^ | South Africa ^c^ |  |  | United States |
| Saudi Arabia ^d^ | United Arab Emirates ^d^ |  |  |  |
| Trinidad and Tobago ^d^ |  |  |  |  |

*Footnote: a= low income, b= lower middle income, c=upper middle income, d= high income*

*Income groups referenced from The World Bank website.* [*https://data.worldbank.org/country*](https://data.worldbank.org/country)  *(accessed on 08/05/2024)*

### Table 4 Quality assessment of the studies included in this review

| **Study ID** | **1. Clarity of the inclusion criteria** | **2. Detailed description of study setting** | **4. Objective, standard criteria used for measurement of the condition** | **7. Valid and reliable outcome measures** | **8. Use of appropriate statistical analysis** |
| --- | --- | --- | --- | --- | --- |
| **Abahussain 2012** | Yes | Yes | Yes | Yes | Yes |
| **Akande-Sholabi 2023** | Yes | Yes | Yes | Yes | Yes |
| **Albaroodi 2019** | Yes | Yes | Yes | Yes | Yes |
| **Alfian 2023** | Yes | Yes | Yes | Yes | Yes |
| **Alghadeer 2021** | Unclear | Unclear | Yes | Yes | Yes |
| **Alqassab 2024** | Yes | Yes | Yes | Yes | Yes |
| **AlRawwad 2021** | Yes | Yes | Yes | Yes | Yes |
| **Babu 2021** | Yes | Yes | Yes | Yes | Yes |
| **Bashatah 2020** | Yes | Yes | Yes | Yes | Yes |
| **Bhayana 2016** | Yes | Yes | Yes | Unclear | Yes |
| **Bungau 2018** | Unclear | Unclear | Yes | Yes | Unclear |
| **Chong 2022** | Yes | Yes | Yes | Yes | Yes |
| **Cole 2016** | Yes | Yes | Yes | Unclear | Yes |
| **Ehrhart 2020** | Yes | Yes | Unclear | Unclear | Yes |
| **Gubae 2023** | Yes | Yes | Yes | Yes | Yes |
| **Gudeta 2020** | Yes | Yes | Unclear | Unclear | Yes |
| **Hagawane 2022** | Yes | Unclear | Yes | Unclear | Unclear |
| **Jankie 2022** | Yes | Yes | Yes | Yes | Yes |
| **Jarvis 2009** | Yes | Unclear | Yes | Yes | Yes |
| **Jha 2021** | Yes | Yes | Yes | No | Yes |
| **Kharaba 2022** | Yes | Yes | Unclear | Unclear | Yes |
| **Low 2023** | Yes | Yes | Yes | Unclear | Yes |
| **Mahlaba 2021** | Yes | Yes | Unclear | Unclear | Yes |
| **McCullagh 2012** | Yes | Yes | Unclear | Unclear | Yes |
| **Michael 2019** | Yes | Yes | Unclear | No | Yes |
| **Michelin 2023** | Yes | Yes | Yes | No | Yes |
| **Nairat 2023** | Yes | Yes | Unclear | Yes | Yes |
| **Painter 2018** | Yes | Yes | Yes | Yes | Yes |
| **Patel 2021** | Yes | Yes | Yes | Unclear | Yes |
| **Purohit 2022** | Yes | Yes | Unclear | Unclear | Yes |
| **Raja 2018** | Yes | Yes | Unclear | Unclear | Yes |
| **Shakib 2022** | Yes | Yes | Yes | Unclear | Yes |
| **Shukla 2023** | Yes | Yes | Yes | Unclear | Yes |
| **Shuleta-Qehaja 2022** | Yes | Yes | Unclear | Unclear | Yes |
| **Srikanth 2023** | Yes | Yes | Unclear | Yes | Yes |
| **Tabash 2016** | Yes | Yes | Unclear | Unclear | Yes |
| **Tai 2016** | Yes | Yes | Yes | Yes | Yes |
