## Supplementary material for "Assessing the knowledge, attitudes and practices of healthcare staff and students regarding disposal of unwanted medications: A systematic review": Abstraction table

| Study characteristics | | Participant characteristics | | Outcome measures | | | | | Current challenges | Future recommendations |
| --- | --- | --- | --- | --- | --- | --- | --- | --- | --- | --- |
| Study (author, year, country) | **Study setting; study design** | **Total number of participants** | **Profession of staff/students and number of participants in each subgroup** | **Knowledge** | **Attitude** | **Advice on the method of disposal** | **Methods of disposal outside healthcare setting** | **Method of disposal within healthcare setting** |  |  |
| Abahussain 2012, Kuwait | 6 government hospitals (secondary) and in the polyclinics (primary); cross-sectional, self reported survey, newly designed by authors | 114 | hospital and community pharmacists | The majority of respondents stated that damage occurs to the environment when unwanted medicines (UMs) are disposed in trash (83%) and when they are disposed of in the sink or toilet (82%). | Majority are willing to have their pharmacies designated as collection site for UMs (86%). 97% feel responsible to protect the environment. | 21 pharmacists refuse taking back UMs because they were unsure of what to do. | not assessed | 72% of pharmacists reported receiving UMs at their work place. 73% in the trash, 32% in sink, 9% in toilet, 20% pass to someone, 10% return to ministry's central drug store, 6% disposed with hospital medical waste. | Absence of policies to accept returned UMs from the public. | Availability of take-back programs. An educational campaign along with proper implementation of the guidelines. Opportunity for pharmacy students to participate in promoting awareness and establishing a national drug take-back program. Students to carry out patient and community education. |
| Akande-Sholabi 2023, Nigeria | a Nigerian university; cross-sectional, self reported survey, derived from previous studies and modified by the authors. | 930 | Medical students (31.4%), Pharmacy students (36.8%), nursing students (10.8%), others (21%) | 62-83% aware improper disposal of UMs can affect the environment and public health. 83.8% reported having no training. 67.7% were unaware of proper disposal methods. | not assessed | 15.5% advised others on how to safely dispose of unused medicines | 67.1% in the trash, 9.5% in toilet, 10.1% return to pharmacy or drug take-back program, 13.3% give to others. 74.2% keep UMs at home. Participants who were aware of the correct disposal method had a higher proportion of good practice (44.0%) compared to those who were not aware (28.3%). | not assessed | Gaps in both training and awareness of proper disposal methods. Lack of access to drug take-back programs and inconvenience. Fear of legal repercussions. | Increase public awareness and education on safe disposal practices. Provide more accessible drug take-back programs. Encouraging healthcare providers to discuss proper disposal with patients. Develop and implement policies. |
| Albaroodi 2019, Iraq | government health sector and private pharmacies at Karbala; cross-sectional, self reported survey, derived from previous studies and modified by the authors. | 129 | hospital and community pharmacists | 46.5% agreed with drug disposal via the trash. 34.9% agreed with drug disposal through the sink and 36.4% via incineration. On the other hand, 65.9% of them agreed to return medication to the source (drug store or company).  58.9% acknowledged the take-back program. |  | not assessed | not assessed | not assessed | Take back program education barrier. | Educational courses on a take-back program for healthcare professionals. Government initiatives for creating awareness for proper disposal. Multi-disciplinary collaboration. |
| Alfian 2023, Indonesia | community health centers; cross-sectional, self reported survey, derived from previous studies and modified by the authors. | 202 | community pharmacists | Half of pharmacists were aware of the environmental hazards of unused and expired household medication disposal (58.4%). | 99% agreed on their responsibility to prevent environmental risks. The majority of pharmacists (80.7%) believed that community pharmacies and pharmacies of private (70.8%) and public hospitals (80.7%) are the most appropriate locations for collecting unused household medications | not assessed | not assessed | not assessed | not stated | Support the initiation of medication take-back programs within community pharmacies. Policies should be developed to govern the collection of returned medications through pharmacies. |
| Alghadeer 2021, Saudi Arabia | community pharmacies; cross-sectional, self reported survey, derived from previous studies. | 360 | community pharmacists | About 80% of CPs reported environmental damage as a result of unused medications being thrown in the sink or toilet. | A very high percentage of CPs (87.5%) agreed that protecting the environment is one of their individual responsibilities. | not assessed | not assessed | Solid: 3.6% trash, 1.1% sink, 1.9% toilet, 16.1% in medicines' bin, 75.3% sent back to pharmaceutical distributor, 1.4% other. Liquid: 1.7% trash, 3.6% sink, 4.4% toilet, 15.5% in medicines' bin, 73.3% pharmaceutical distributor, 1.1% other. | Unavailability of policies/ guidelines. | Implement take-back programs. A policy to be formed by the Ministry of Health, which would permit community pharmacists to receive returned UMs from the general population and guide the pharmacists on their appropriate disposal. |
| Alqassab 2024, Saudi Arabia | from tertiary care hospitals to smaller community clinics; cross-sectional, self reported survey, derived from previous studies. | 321 | pharmacists (21%), nurses (22%), physicians (27%), others (30%) | Low level of staff awareness of the medication waste issue. 59% strongly agreed that inappropriate disposal of medication waste could result in harm for the environment. | 60% seek for advice during their work life. 57% strongly agreed that medication waste should be an issue for each person handling medications. 95 % showed a willingness to practice safe disposal of medication waste. | not assessed | not assessed | 42% in the trash, 23% in specialised medication waste container, 23% in yellow containers | Issues with availability of collection bins, accessibility. Limited knowl­edge about the issues. Lack of specific guidelines and overprescribing. | Appropriate lo­gistic (bins, containers, accessibility) to promote proper medication waste disposal. Policies from governmental bodies. Training and work­ shops to highlight the importance of safe disposal of medication waste. Promoting interprofessional collaboration in the context of medication waste management. |
| Al Rawwad 2021, United States | a public university in Houston; cross-sectional, self reported survey, derived from previous studies. | 210 | pharmacy students (66%), nursing students (34%) | 47% pharmacy students and 68.9% nursing students did not receive advice on proper disposal of medications. 79.4% pharmacy and 80.4% nursing students chose a medication bin collected by a contractor as the most appropriate method to dispose of unused medicines. 11.8% pharmacy and 34.6% nursing students were not sure of how to dispose medicines. | 36.8% nursing students reported nurses as the main provider of advice on safe disposal of medicines. 82% pharmacy students reported pharmacists as their main provider of advice on safe drug disposal. | not assessed | Pharmacy and nursing students respectively: Solid: 33.1%, 19.7% in trash; 0%, 1.4% sink; 2.9%, 5.6% toilet; 82%, 85.9% in medicines' bin collected by contractors; 42.4%, 40.8% pharmaceutical distributor; 5%, 5.6% other. Liquid: 25.9%, 14.1% in trash; 11.5%, 19.7% sink; 6.5%, 12.7% toilet; 80.6%, 77.5% in medicines' bin' 41.7%, 36.6% pharmaceutical distributor; 3.6%, 5.6% other. | not assessed | Lack of advice from healthcare professionals. | One future educational strat­egy would be the incorporation of simulation and role play as an interactive unfolding case study for an inter-professional educational endeavor. |
| Babu 2021, India | tertiary care university hospital pharmacies and community individual or chain pharmacies in Chennai city; cross-sectional, self reported survey, derived from previous studies and modified by the authors. | 172 | hospital pharmacists (HP) (50%), community pharmacists (CP) (50%) | 62% HP stated returning UMs for all dosage forms to the source. Whereas 52% CP chose trash for solid dosage forms; 48% chose sink for liquid; 60% return to the source for parenteral and semi-solid. 95% of the HP and 81% of the CP were aware that improper disposal of drugs could be a cause for environmental pollution. Awareness of guidelines: 85% HP, 62% CP. Awareness of take-back system: 76% HP, 57% CP. | Less than 5% HP and 10% CP had a negative or lack of opinion on the impact of improper disposal of drugs on environmental pollution. Both hospital (84%) and community pharmacists (91%) consented to participate in the educational programs focusing on take-back of drugs and safe disposal of expired and unsafe medications. | Majority of the pharmacists agreed that they had never given advice to customers about how to dispose unwanted medicines.  65% of HP had accepted that they had displayed posters that explain the safe disposal of drugs in their pharmacies, while 95% of the CP did not have any posters in their pharmacies. | not assessed | 88% of hospital pharmacists stated that the collection boxes were available at the hospital pharmacy. On the contrary, 74% of community pharmacists said that there were no collection boxes at community pharmacies. | Most of the community pharmacists were not aware of and were not practicing the collection boxes. Even though few community pharmacies have collection boxes, there was no utilization of them. The drug take-back program is not functional. | Keep collection boxes for expired and unused drugs in pharmacies. Poster display in pharmacies in educating people on the safe disposal of the drugs. The statutory bodies to create awareness among the pharmacists working in community settings through continuing educational programs and newsletters focusing on safe medication use and disposal practices and legislations for the development of drug take-back program. |
| Bashatah 2020, Saudi Arabia | university; cross-sectional, self reported survey, derived from previous studies. | 352 | pharmacy students (46%), nursing students (54%) | 81.8% pharmacy and 91.9% nursing students accepted that inappropriate disposal of unused medicines can affect the environment and health. | 19.9% pharmacy and 16.3% nursing students agreed that the responsibility for creating awareness of proper disposal of UMs lies with pharmacists. 64.6% pharmacy and 81.6% nursing students agreed responsibility lies with the Ministry of Health. | not assessed | Pharmacy and nursing students respectively: 47.2%, 61.2% trash; 6.8%, 5.3% sink or toilet; 6.8%, 4.7% medical store or pharmacy; 37%, 52% stored leftover meds at home; 15.5%, 20.5% give to friends/relatives. | not assessed | not stated | Increase education among health care students and the general public. This can be accomplished by the government, learning institutions, and in pharmacies at the point of sale. Avoid overprescribing. Recommend the Ministry of Health issue official guidelines and policies for the safe disposal of UMs. The government should establish a take-back program in partnership with medical stores and the pharmaceutical industry. |
| Bhayana 2016, India | tertiary care public hospitals, private pharmacies; cross-sectional, self reported survey | 300 | pharmacists (33%), doctors (33%), nurses (33%) | not assessed | 95% doctors, 94% nurses, 66% pharmacists believed that it is everyone's responsibility to dispose unused medicines. 84% pharmacists expressed their concern toward protecting the environment from disposed medicines. | not assessed | 76% nurses, 59% doctors, 70% pharmacists disguise and dispose medicines in trash. 8% nurses, 16% doctors, 8% pharmacists flush in toilets. | Only 8% of the HCPs received unused drugs back from consumers. 59% nurses, 37% pharmacists, 47% doctors dispose unused medicines in correct containers. | inadequate awareness and practices | Design an innovative policy for the drug take‑back program, and to motivate the health regulators to strengthen and implement the existing drug disposal policies more effectively. Teaching undergraduate and postgraduate medical, nursing, and pharmacy students and conducting continuous medical education training for healthcare professionals on the use, collection, and disposal of unwanted drugs. |
| Bungau 2018, Romania | independent pharmacies and chain of pharmacies in urban and rural area; cross-sectional, survey done by phone and on-line | 521 | pharmacists (79%), pharmacy assistants (21%) | More than half of the pharmacists (53.6%) consider themselves sufficiently informed about the waste disposal legislation, only 19.3% consider themselves highly informed. | 75% stated they are responsible collecting unused medicines from the citizens. Over 65% were dissatisfied with the current procedure. 68.7% consider it extremely important to inform patients about the correct way to disposed unused medicines. | not assessed | not assessed | 16% do not collect unused medicines from the population. About 45% said that, in the pharmacy where they work, less than 25 kg of waste are collected per year. Only 2% of the pharmacies that collect medical waste are required for this service daily. | A lack of procedure, incomplete legislation, exceeding the amount contracted with the operators, and high costs of disposal service. Population’s low interest, poor information, and increased drug use. | Funding support by patients, local authorities, manufacturer, pharmacies. A clear and easy procedure. Suggest the placement in pharmacies of special containers where citizens can directly put unused medicines. More information campaigns. Decrease the quantity of prescribed medicines. The implementation of a unitary system at the national level to increase efficiency of the service. Pharmacists’ representatives to be directly involved in the drafting of legislation. |
| Chong 2022, Malaysia | community pharmacies; cross-sectional qualitative study, telephone semi-structured interview | 18 | community pharmacists | not assessed | 78% willing to provide medicines return services. All feel responsible to support government initiatives. All respondents agreed that safe medicine disposal is beneficial for public health and environment. 56% considered environmental impact as the reasons for their readiness to provide medicines return services. | 89% did not accept medicines return from customers. Some respondents suggested that customers utilise the Medication Return Program (MRP) available at government healthcare facilities, while some did not suggest any alternative safe medicine disposal methods. | not assessed | not assessed | No formal guidelines or requirements for community pharmacies. Lack of service availability. Lack of demand for such services from their customers and lack of public awareness. High operational costs for the pharmacies. | Subsidies from the government and consumers to help with the extra costs of the medicine return service. Providing a convenient location for consumers to dispose of unwanted medicines. Collaboration with non-profit organisations. Provide guidelines for community pharmacies. |
| Cole 2016, United States | Universities in Florida; cross-sectional, self reported survey | 278 | pharmacy students | Over 30% were unaware of proper medicines disposal. 67% were aware of where their local drug take-back sites are located. 65% reported that they had received exposure to medication disposal in the classroom curriculum. | 40% of participants reported a score of 5 or less on the confidence scale indicating they were not very confident in drug disposal. 5% of students reported that they were extremely confident. | Solid: 37% unsure, 9.4% disposal site, 4.4% trash, 43% mixed garbage, 3.3% sink/toilet, 0.8% sealed bag, 1.9% bring to pharmacy. Liquid: 44.3% unsure, 8.9% disposal site, 3.2% trash, 27.6% mixed garbage, 12.4% sink/toilet, 1.4% sealed bag, 2.2% bring to pharmacy. | not assessed | not assessed | Lack of awareness of proper medication disposal due to lack of standard education, lack of reintroduction or application within curriculums, or lack of student interest or appreciation of importance. | Early introduction and repetitive application throughout pharmacy curriculums, more standard education resources provided to each curriculum. Suggest various teaching methods including a dispensing lab or objective structured clinical examination (OSCE). Emphasize the pharmacist's responsibility for proper medication disposal in order to improve patient and environmental outcomes. |
| Ehrhart 2020, United States | community pharmacies in Portland and Oregon, eight with a dropbox and 17 without a dropbox; cross-sectional qualitative study, interview | 29 | community pharmacists | not assessed | 65.5% expressed positive attitudes regarding drug take-back programs. All other pharmacists except one at dropbox locations expressed positive attitudes towards take-back programs. | Pharmacists at dropbox locations recommended primarily to use the onsite dropbox, and never told customers to flush or throw away drugs. Many pharmacists redirected customers to look elsewhere, such as the DEA website or their local garbage collector for the disposal method. | Pharmacists with dropbox locations: unsafe disposal (e.g. store at home, trash, flush) 78.7%; safe disposal (e.g. dropbox, pharmacy) 21.3%; Pharmacists without dropbox locations: unsafe disposal 89%; safe disposal 9.5%. | not assessed | Lack of legislative efforts on environmental issues. Hinderances to establishment and use of dropboxes e.g. cost, lack of public education/awareness, and liability for the pharmacy. Lack of floor space for small stores. Policy inconsistencies in terms of variability in drug types and forms permissible for collection at different dropboxes. | Legislation to include environmental perspective. Establish dropboxes in all pharmacies. Pamphlets, brochures, and stickers on medication bottles as options to deliver information to customers. Funding support from government grants, customer fees, and the pharmaceutical industry. Changes to online information from federal and local agencies to improve consistency. Development of pharmacy school and employment training programs. |
| Gubae 2023, Ethiopia | four universities in Northwestern Ethiopia; cross-sectional, self reported survey, derived from previous studies and modified by the authors. | 445 | pharmacy students | 27% had been taught about proper disposal of medicines in pharmacy school or by health professionals. 60% stated improper disposal of medicines contributes to environmental pollution. 20.4% aware what ecopharmacology is. 41.6% aware of guidelines. | 87% supported the implementation of policies to safely dispose of medications. 79% were willing to dispose of medications in an appropriate place. 82.5% agree that manufacturers and pharmacies should collect UMs from the public. 90% agree that pharmacists are responsible for protecting the environment from pharmaceutical waste. | not assessed | 32.4% in toilet/sink; 21.8% give to friends or relatives; 61.8% in trash; 20.2% burn; 6.1% returned to pharmacy/ hospital. 63.8% stored unwanted medicines at home. | not assessed | Lack of education about medication disposal. | Aware of the life cycle of a drug. More uniform education on ecopharmacology in pharmacy education and mandatory teaching of drug disposal. Government should consider investing in pharmaceutical take-back programs. |
| Gudeta 2020, Ethiopia | private retail pharmacies and drug shops, Jimma city; cross-sectional, self reported survey | 87 | private pharmacists (84%), nurses (10%), others (6%) | 61% had awareness regarding appropriate disposal practices and safe disposal sites. 41.7% knew that safe disposal of expired medicines would prevent environmental pollution.18.4% received training on pharmaceutical waste management. | 74.7% strongly agreed that improper disposal would negatively affect health and ecological systems. 59.8% strongly agreed that environmental protection is their responsibility. 46.9% and 38.5% of participants respectively held the pharmaceutical supply agency and government authority accountable in creating awareness for the drug retail outlets. | not assessed | not assessed | 38.2% burning separately at retail outlets, 19.5% burying underground, 14.6% store in quarantine until received by district health bureau, 13.8% flushing in toilet or rivers, 8.9% in trash, 4.1% return to suppliers, 2.4% transfer them to other retail outlets. | Infrequent inspection of retail outlets by the regulatory body, where most of them received once a year. Apart from the curricular programs, it is not common for the government to provide short-term training for private practitioners. | Government should increase the frequency of inspections and also raise practitioners’ awareness of safe disposal practices. Government should consider private practitioners during the provision of training on healthcare waste management for public health professionals. |
| Hagawane 2022, India | community pharmacies in urban area; cross-sectional, self reported survey, predesigned | 133 | community pharmacists | 75% were not aware of the existence of guidelines. 62.5% sent expired medicines to the distributor, but unaware what happens to the expired medicines after returning. 12.5% thought that distributor might reuse the medication. | not assessed | not assessed | not assessed | On observing the disposal method of expired medicine, the majority of the pharmacist returns it to the distributor (81.3%). Majority of solid expired medicine were returned to the distributor (75%) similar to this for liquid medicine 73.4 %, remaining of them uses other methods of disposal like throwing in the dustbin, burning, dumping or disposing of in drainage system. | Unawareness of guidelines due to improper education about expired medicine disposal in their pharmacy education. | Providing adequate training for pharmaceutical waste management to all the pharmacy graduates. The guidelines regarding pharmaceutical waste disposal should be implemented and supervised more strictly by controlling authorities. |
| Jankie 2022, Trinidad and Tobago | public and private (community) sectors in Trinidad; cross-sectional, self reported survey, derived from previous studies. | 208 | public and private sector pharmacists | 64% would not recommend flushing meds down the toilet. 40% incorrectly stated that OTC medicines can be disposed in household trash. 79.3% correctly identified that improper disposal of pharmaceuticals can have negative implications for planetary health. | A large portion (79.8%) thoughts that pharmacists should be the source of drug disposal information, although 64.5% believed that the Ministry of Health should be responsible for the mass dissemination of drug disposal information. Just 28.4% believed that it is a part of the pharmacist's duty to provide drug disposal information to patients. 69.2% would like their workplace to be a medication takeback site. | 20.3% private, and 72.3% public pharmacists give advice on medication disposal. Only 38% reported patients asking for drug disposal advice. | not assessed | 80% return to pharmaceutical distributor, 63.9% incinerate through drug inspectorate, 32.3% dispose in workplace garbage. | The state disposal service is only available to facilities and not to patients, but community (private) pharmacies can access the service for a fee whilst it is provided free of charge to state-run facilities. No medication take-back programme implemented in Trinidad. Minimal training and knowledge in proper medication disposal. | Increase campaigns by the Ministry of Health to educate the population on good disposal practices. The need for training and continuing medical education for pharmacists including adequate disposal information. The need of a national policy for safe medicines disposal. |
| Jarvis 2009, United States | Hospital or community practice; cross-sectional, self reported survey, pre- and post- educational intervention | 158 | hospital and community pharmacists | 36% stated never learned about proper disposal. Knowledge pre- and post- education intervention in the form of a newsletter. Pharmacists perceived inappropriate medicines disposal to be an environmental problem: pre-intervention 47%, post 57%. Disposed medicines down the sink: pre 19%, post 5.6%. Flush to toilet: pre 27%, post 22%, In trash: pre 13%, post 16%. Return to pharmacy: pre 21.35%, post 25.75%. Arranged for hazardous waste pick up: pre 10%, post 20. | Pharmacists are good resources for information regarding proper disposal: strongly agree presurvey 41.6%, postsurvey 46% | Pharmacist recommendation for unused medication pre- and post-intervention respectively: Dispose in trash (12.76%, 15.67%); return to pharmacy (21.35%, 25.75%); flush down toilet (27.08%, 22.01%); wash down sink (19.01%, 5.6%); arrange for hazardous waste pick up (9.9%, 20.15%); other (9.9%, 10.82%). | not assessed | not assessed | not stated | Incorporating proper medication disposal practices into pharmacy education curricula and continuing education programs. Increase pharmacists' awareness of proper medication disposal especially in light of the new federal recommendations designed to protect patients and the environment. |
| Jha 2021, Nepal | KIST Medical College and Teaching Hospital; cross-sectional, self reported survey, derived from previous studies and modified by the authors. | 441 | undergraduate medical students (70%), other (undergraduate dental students) (30%) | The knowledge median score was 8 out of 10. | not assessed | Over 40% of respondents had educated their friends and family members about the safe disposal of medicines. | 38.5% in trash, 4.1% toilet, 44.7% kept at home, 3.4% burn with garbage, 9.3% return to pharmacy shop. | not assessed | No safe medicine disposal guidelines adopted in community pharmacies. Knowledge about drug takeback system is low and it is not widely adopted in the country. Challenge of bringing the medicines back to the pharmacy from where it was purchased and of providing the proper documentation. | More aware­ness and creating a chart of disposal procedures of common medicines and popularizing the chart can be important to strengthen knowledge. The drug takeback system should be implemented in community pharmacies initially located in major cities and information about the system should be widely disseminated. |
| Kharaba 2022, United Arab Emirates | Community pharmacies; cross-sectional, self reported survey, derived from previous studies and modified by the authors. | 418 | community pharmacists | Knowledge of the ways to destroy collected expired medicines: 31% incineration, 20% general garbage; 10% toilet; 7.2% sink; 32% not sure. | 68.4% think UAE need a specialised center for medication disposal. The funding of this center should be by: Ministry of Health (64.7%), patients (11.2%), pharmaceutical companies (13.3%), community pharmacists (10.8%) | not assessed | not assessed | Solid: 39.5% contractors, 14.9% trash, 10.2% sink, 9.1% toilet, 26.4% return to distributor. Semi-solid: 37.1% contractors, 11.5% trash, 11.5% sink, 14% toilet, 25.9% distributor. Liquid: 39.1% contractors, 8.9% trash, 9.3% sink, 14.9% toilet, 27.8% distributor. | Cost implication for returning medicines to contractors. Distributors only took back what they had previously agreed upon with the pharmacists during the sale. Education gap. Lack of system or guidelines. | Action to manage the quantities of expired medications: stock limitation, collaboration with other pharmacies to exchange the nearly expired drugs. The establishment of specialised center for medicines disposal in UAE. |
| Low 2023, Malaysia | Malaysian community pharmacy guild event; cross-sectional, self reported survey, designed by authors | 168 | community pharmacists | Mean knowledge score 6 out of 10. 43.5% CP have not received education concerning medicines waste disposal. 85.7% aware medicines disposed in landfills are harmful for the environment. 21.4% answered correctly on question relate to Medication return programme. | 65% were somewhat confident to provide advice related to medication disposal. 87% think it's their responsibility to ensure correct medicines disposal. 94% believe that medication return programme (MRP) should be set up in community pharmacies. 94% think the impact of medicines on the environment is important topic. 97% are motivated in encouraging patients to dispose their medicines appropriately. 94% said it's their responsibility to educate patient. 99% would like training on proper disposal of medicines. | Respondents advising patients: MRP 79.8%. 0.6% toilet, 2.4% trash, 13% not asked, 4.2% other advice. 79.2% CP were never asked or asked <5 times/year by patients for medicines waste related questions. | not assessed | not assessed | Education gap. Lack of proactive efforts by community pharmacists in promoting safe medicines disposal despite satisfactory knowledge and attitude. Lack of funding and legislative support for medication take-back programs. | Funding from Ministry of Health towards MRP in community pharmacies. More proactive and contributory efforts from community pharmacists, supported by public and private sectors to bridge the gap between knowledge and practice. |
| Mahlaba 2021, South Africa | Sixteen randomly selected primary health-care clinics in two subdistricts of the City of Tshwane, Gauteng Province; cross-sectional, self reported survey, derived from previous studies. | 165 | Pharmacists (1.2%), Pharmacist assistant (11.4%), medical practitioner (5.4%), nurses (80%), dental (1.8%) | 90% were aware of waste disposal SOPs. 25.6% were not aware of proper medicines disposal. 71% said they did not have formal or informal training received on safe disposal of medicines. | not assessed | 71% always or sometimes counsel patients regarding safe disposal of medicines. 65% HCPs have never been asked by patients. Information provided to patients by HCPs: 22% return medicines back to the clinic, 11% only taught about segregation of medicines, 5.6% disposed UMs into drain. | not assessed | 76.5% HCPs never participate in the destruction of medicines. Disposal methods: Incineration 31.9%, Landfill 4.8%, flush down the toilet 20.5%, flush down the basin 9.6%, dissolve in boiling water 8.4%, municipal waste 7.8%, burning it at the clinic 4.8%, unsure 9.6% | Lack of knowledge by the healthcare professionals on proper disposal methods. Lack of patients counselling by healthcare professionals about the safe disposal of medicines despite agreed SOPs. | More innovative educational strategies to improve HCPs' knowledge. e.g. greater internet use. More frequent reviews of national and regional government disposal protocols with guidance actively disseminated. Media (TV and radio) can also be used to encourage patients to safely dispose of their unwanted medicines through HCPs and other trusted sources. Suggesting proper medicines disposal topic to be included in the curriculum and ongoing professional development. |
| McCullagh 2012, United States | Hospice home care settings in Michigan; cross-sectional, self reported survey, designed based on earlier survey | 138 | hospice homecare nurses | Respondents most often indicated that their sources of information about drug disposal practices came from the hospice manual, rules, other documents, or hospice inservice (62% each). Nearly one-third of nurses reported that they were not at all or only a little familiar with the rules on medication disposal. | Most hospice nurses reported they were moderately or extremely concerned about legal and environmental issues, but diversion of drugs was the highest level of concern. | not assessed | Always or often: Flush 37%, rinse 18%, pharmacy 2%, hospice 1%, mix with noxious 64%. Sometimes or never: Flush 63%, rinse 82%, pharmacy 98%, hospice 99%, mix with noxious 36%. | not assessed | A lack of alignment with federal guidelines for drug disposal. Lack of awareness of guidance. | Further education, practice, and research directions, emphasizing the need for hospice nurses to be more aligned with federal guidelines for environmentally safe drug disposal. |
| Michael 2019, Nigeria | Community pharmacies registered across Anambra State, Nigeria; cross-sectional, mixed method with survey derived from previous study and interview | 77 | community pharmacists | not assessed | 71.4% the need of a state-run medicines disposal scheme. Respondents want to have lectures on pharmaceutical waste management to be incorporated in schools of pharmacy curriculum (100%). They agree there is a need for improvement in the disposal practice (100%). | not assessed | not assessed | Solid: 23.9% in trash, 0% sink/toilet, 35% NAFDAC, 32% drug wholesalers/ distributors, 9% burn. Liquid: 22.4% trash, 7.1% sink, 0% toilet, 31.8% NAFDAC, 34.1% wholesalers, 4.7% burn. Class B controlled drug: 9.9% trash, 0% sink, 1.2% toilet, 29.6% NAFDAC, 24.7% wholesalers, 4.9% burn. 54.5% did not comply with NAFDAC guideline on disposal of expired drugs. | Poor education, awareness and documentaries on management of expired drugs. Poor law enforcement. Stress, and inadequate protocols involved in returning medications to government agency. Policy implementation gap between the national government and the participants. | Suggest NAFDAC to fund the state-run medicines disposal system that will augment the federal system. Create public awareness on the consequence of improper management of expired medicines. Adequate training of health workers on the management of expired drugs. Implementation of a local government- run disposal system. Adequate law enforcement strategies. Initiate programs that will make standard disposal practice easier and less time-consuming. |
| Michelin 2023, Brazil | Community pharmacies in the State of São Paulo, Brazil; cross-sectional, self reported survey, derived from previous studies. | 630 | community pharmacists | 58% aware of Brazilian legislation on the reverse logistics of unused household medicines. 42% said they have received training on reverse logistics for medicines. 97% think the improper disposal of medicines affect the environment and human and animal health. | 59% think pharmacists should provide guidance to customers on disposal of unused medicines. 50% think nurses, 20% think doctors. | Respondents advising patients: 1.4% in trash, 0.78% toilet, 80% return to pharmacy, 17.5% other. 80% customers rarely or never ask for guidance on disposal of medicines. | not assessed | 68% has establishment to accept unused medicines for disposal. | Lack of customer inquiries about proper disposal methods implies a potential awareness gap. Education gap. | The need for professional training and further education programs for pharmacists. A need for policies and practices that minimize environmental harm from medication disposal. |
| Nairat 2023, Palestine | Community pharmacies across the West Bank, Palestine; cross-sectional, self reported survey, derived from previous studies. | 400 | community pharmacists | 87% agree improper drug disposal can cause environmental harm. | 97% think country needs a national drug disposal system. Majority (78%) think the district health board should fund drug disposal system. 94% think it's their responsibility to protect the environment. | Solid drugs: 63% trash, 3.5% toilet, 0.3% sink, 2.8% incineration, 19.5% return to pharmacy, 4% donation to hospitals, 1.5% giving to friends/family, 4.8% none recommended, 0.8% other. Liquid: 32.8% trash, 10.8% toilet, 33% sink, 0.8% incineration, 12.5% return to pharmacy, 1.8% donate to hospitals, 1.5% give to friends/family, 5.8% none recommended, 1.3% other. | not assessed | Methods of disposal of UMs in pharmacy: 48.3% via trash, 3.5% via toilet, 22.5% sink, 5.8% incineration, 35.3% drug take-back box, 73.3% return to companies, 20.8% return to Ministry of Health, 1.8% other. | Poor education and awareness about the management of unwanted drugs. Poor law enforcement, difficulty returning drugs to disposal site due to lack of formal policy, lack of documentaries on the management of unwanted drugs. difficulty of accessing drug drop-off sites. | Establishment of a national drug disposal system with a preference for funding by district health board. Law implementation on safe drug disposal. Organising events, distribute brochure to encourage pharmacists to stay up to date on the topic continually. Provide proper training to healthcare workers. |
| Painter 2018, United States | Community pharmacies in San Diego County, California; cross-sectional, self reported survey, derived from previous studies. | 36 | community pharmacists | 75% aware that Drug Enforcement Agency had recommendations regarding medication disposal. Pharmacists received information about medication disposal primarily through work training (72%) and government agencies or professional organizations (58%). Pharmacists were aware that medications could be disposed at National Drug Take-Back events (83%). | 100% felt comfortable providing medication disposal education to their patients. 100% think it's important that pharmacists provide education on medication disposal. | 83% National Drug Take Back events for non-controlled drugs, 89% controlled substances. 8% recommend flush down the toilet or sink. 6% recommend return back to pharmacy. 42% recommend non-controlled substances to be mixed with undesirable substances and dispose via trash, 22% recommended it for controlled substances. | not assessed | not assessed | Lack of engagement in disposal education due to increased workload. Inconsistencies in guidelines. Lack of comprehensive knowledge: while most pharmacists were aware of the DEA recommendations, there was less awareness about FDA, EPA, and local county agencies' guidelines. Lack of disposal education through pharmacy school. | Encouraging pharmacists to ask their patients about medication disposal or incorporating this information into consultation could help create opportunities to provide this education more frequently and establish pharmacists as a resource for medication disposal information. The need for strengthening education on medication disposal in pharmacy school curricula and for programs or continuing education to keep pharmacists up to date on regulations and proper disposal methods. |
| Patel 2021, India | A tertiary care teaching hospital in Ahmedabad, India; cross-sectional, pre-validated investigator-administered questionnaire | 170 | doctors (45.9%), nurses (49.4%), pharmacists (4.7%) | 15.8% recognised returning to donor or manufacturer as a safe disposal method, 37.6% considered landfilling safe, 11.1% incineration, 11.1% chemical decomposition, 24% all of above methods. 54% unsure if medicines can be filtered out during water sewage treatment. | 74% agree or strongly agree that disposal of medicines is responsibility of law enforcement agency and healthcare system. 78% agree or strongly agree that UMs which are disposed via toilet, sink or trash have harmful impact to environment. 53% believed there is no unused medicines disposal policy in place in India. | not assessed | 66% sometimes, usually or always dispose liquid into sewer.  15.9% sink, 31% toilet, 35.6% trash, 8.3% public place, 5.3% river, 2.2% well. | not assessed | Lack of uniform knowledge or guidelines. Absence of formal drug disposal policy and program. | The need for an effective drug disposal policy and program. Adequate training programmes. |
| Purohit 2022, India | Pharmacies dispensing drugs to patients, including those situated in hospitals, within the vicinity of All India Institute of Medical Sciences Rishikesh, Uttarakhand, and the city of Dehradun, Uttarakhand; cross-sectional, structured questionnaire | 373 | owners of pharmacies and hospital pharmacies | Knowledge regarding disposal of different dosage forms. Solid dosage form: 88% return to manufacturer, 0.5% throw in dustbin with intact packaging, 1.6% throw in dustbin without packaging, 0% flush in toilet, 6.14% burn in open containers, 3.5% high temperature incineration. | 95.7% would like to learn more about safe disposal of expired medicines. There is a consensus that proper disposal of expired medicines is primarily the responsibility of pharmacists (72.6%), with some also believing it to be a collective responsibility involving nursing professionals (1.88%) and municipal corporations (1.07%). | not assessed | not assessed | Solid: 88.2% return to manufacturer, 0.5% throw in dustbin with intact packaging, 1.6% throw in dustbin without packaging, 0% flush in toilet, 6.14% burn in open containers, 3.5% high temperature incineration. Parenteral: 95.4% return to manufacturer, 0.3% dustbin with intact packaging, 1.9% dustbin without packaging, 0% toile, 1.1% burn in open containers, 1.3% high temp incineration. Controlled drugs: 99.7% return to manufacturer, 0.3% burn in open containers. | Lack of well defined guidelines and lack of policy regarding the disposal of expired medicines in India | Conduct lectures for all pharmacists, and in local area, newsletters, workshop and site visits, periodic emails and internet-based updates. The need for innovative policy implementation by health regulators to strengthen drug disposal practice. |
| Raja 2018, India | SRM Medical College Hospital & Research Centre, Chennai, Tamil Nadu, India; cross-sectional, semi-structured questionnaire | 393 | doctors, medical students, pharmacy students, and nurses | Only 11.1% of participants had partial knowledge about proper drug disposal practices. A significant portion of participants (89%) recognized that improper disposal could lead to environmental pollution. | not assessed | 26% always advised for proper drug disposal, 43% advised occasionally and 31% never advised. | 17% throw in trash, 17% crush the drug before discarding, 6% flush in toilets or sinks, only 5% return to medical store. | not assessed | A lack of utilization of official drug take-back programs or formal guidelines for disposal. | Implement national drug take back programme by creating awareness through government programmes and via healthcare professionals. Creating robust systems that not only facilitate but also enforce the proper disposal of medications to prevent environmental pollution. Utilisation of social media and other platforms to create awareness. |
| Shakib 2022, Bangladesh | Top 10 private universities in Bangladesh; cross-sectional, structured questionnaire, derived from previous studies. | 250 | pharmacy students (60%), general students (40%) | 63% aware of environmental hazard due to improper disposal of medicines. 64% unaware of the standard drug disposal method. 87% said they do not have drug take-back system in their society. | 80% encouraged recruiting pharmacists to counsel proper medicines disposal. | not assessed | Solid: 85.6% in trash, 4.8% follow standard disposal method, 6.4% return to pharmacy, 3.2% sink and toilet. Liquid: 64% trash, 10% follow standard disposal method, 4% return to pharmacy, 22% sink/toilet. 65% pharmacy students store unused medicines at home. | not assessed | Absence of official guidelines for drug disposal and low awareness among the students. | Establish a drug take-back system. Increase awareness about the hazardous effect of unused medicines. Recommend TV and internet as the best source of awareness. Organise seminars and workshops by government-regulated drug administration, environmental scientists, physicians, and pharmacists to raise community awareness. Training for healthcare practitioners and community pharmacists to educate patients. |
| Shukla 2023, India | University Dr. Ram Manohar Lohia Institute of Medical Sciences, Lucknow, India; cross-sectional, self reported survey | 200 | medical students | 52.5% follow the protocols for disposal of medicines. 78% aware of drug take-back program. | 95% think there is a need of systemic program for collection of expired drugs by government or manufactures. 90.5% of the students agree that having a medical background makes them more concerned about the environmental impact of unused medicines. | not assessed | 65.5% in trash, 2% toilet/sink, 5% stored at home, 16.5% return to medical stores, 11% giving to municipal corporation. | not assessed | Students unaware of the proper ways to disposal of medicines with no special protocol in place. Lack of systemic program for collection of expired drugs and lack of public awareness. | Information through newspapers or TV. Written instructions on packets of drugs. Information by doctors, pharmacists, and nurse. Information by village health workers. Establish take-back program by government or manufacturers. Incentives for returning unused medicines to pharmacies. Establish national guideline and publicise. Encourage pharmacist involvement in medication disposal education. Encourage rational prescribing practices, improving medication adherence. |
| Shuleta-Qehaja 2022, Kosovo | A medical college in Kosovo; cross-sectional, self reported survey, derived from previous studies and modified by the authors. | 336 | pharmacy students (49.7%), nursing students (50.3%) | 95% pharmacy and 97% nursing students are aware of environmental damage from inappropriate disposal of medicines. | Responsibility for disposal of unused medicines: Pharmacists: pharmacy students (30%), nursing (36%); The Ministry of Health: pharmacy (64%), nursing (68%); Ministry o the Environment and Spatial Planning: pharmacy (36%), nursing (17%) | not assessed | 78.4% pharmacy and 74% nursing students have unused medicines at home. Pharmacy students 52% in trash, 9% in sink/toilet, 11% return to pharmacy, 16% give to friends/ relatives. Nursing students 56% in trash, 6% in sink/toilet, 11% return to pharmacy, 21% give to friends/relatives. | not assessed | Education gap. Unused medicines as per current legislation can be returned only to pharmacies where the patient bought the medicine by presenting the invoice, which might cause impracticalities for patients. | Organise training, seminars and workshops for health professionals especially for pharmacists and nurses, since they pass information to patients/ consumers. Amendment of the regulations to avoid additional administrative burdens. Support from international institution whose programs are directed towards the environment and climate change. |
| Srikanth 2023, India | A medical college in South India; cross-sectional, self reported survey validated by subject experts | 118 | medical students | 83% aware of medicinal waste. 61.8% know about the safe disposal of unused, expired medicines. 93% agreed that unsafe disposal of unused medicines would adversely affect the environment and human health. | 66% agree there is an inadequate information regarding safe disposal of medicines. 59.3% said that pharmacist is the most appropriate person to provide information. Few said doctors (21.1%), nurses (13.5%), media (5.9%). 58% agreed that public and students should be educated regarding harmful effects of medicines disposal on environment. 58% strongly agreed that educational activities like take-back programs should be introduced. | not assessed | 66% kept leftover medicine at home. 73% trash, 3% give away to friends/relatives, 7% sink, 6% toilet, 7% return to pharmacy, 4% others. | not assessed | Despite awareness of the harmful impacts of improper drug disposal, a significant portion of students lacked knowledge of safer disposal methods, resulting in the following of inappropriate disposal practices at home. | Establish national-level guidelines and conduct awareness programs mediated through healthcare professionals to encourage proper disposal of unused or expired medications. |
| Tabash 2016, Palestine | Five governmental hospitals in Gaza Strip: European-Gaza, Nasser, Al-Aqsa, Al-Shifa, and Kamal Odwan Hospitals; Pre/post-test intervention, cross-sectional, interviewing survey, derived from international guidelines and modified by the authors. | 530 | pharmacists 8.7%, nurses 68.7%, others (radiographer, waste worker) 22.6% (Total number of participants Pre-intervention phase: 530 health care staff. Intervention and post-intervention phases: A subsample of 69 individuals.) | Pharmaceutical waste cause environmental hazard: pre-intervention 83%, post- 94%.  Steps of pharmaceutical waste management: pre- 47.5%, post- 71.9%.  Mean knowledge score: pretest 48%, post- 68%, follow up 81%. | not assessed | not assessed | not assessed | Pre-intervention: the majority of health care staff's practices were rated as poor (44.4%) and fair (55.6%) practice. Post-intervention: satisfactory practices reported by 64.7% of the participants, and poor practices dropping to 5.9%. The intervention led to better segregation of hazardous pharmaceutical waste, proper use of designated containers for waste collection, and adherence to safety protocols during disposal. Mean practice score: pretest 34.3%, post- 44.2%, follow up 78.3%. | Knowledge gap and awareness gap among healthcare staff. Absence of standard guidelines and training. Insufficient resources to facilitate proper disposal practices: color-coded bins and designated areas for safe waste storage. | Educational initiatives could effectively enhance pharmaceutical waste management practices among healthcare professionals. Regular training workshops and the provision of necessary resources for drug disposals. |
| Tai 2016, United States | Community pharmacies in California, USA; cross-sectional, self reported survey | 142 | community pharmacists | 15.9% select appropriate recommendation for disposal of non-controlled drugs; 10.1% controlled drugs. 13.4% aware that all 3 organisation (DEA, EPA, FDA) were information sources for proper medication disposal. Pharmacists were generally aware of drug take-back events as an appropriate method of disposal for medications (72.5%). | Mean attitude score was 16.1 out of 20. Most respondents felt comfortable providing medication disposal education (90.8%) and considered it an important (99.3%) and valuable opportunity (98.6%) to contribute to their patients. Over 80% of the respondents believed that patients and health professionals would like pharmacists to provide medication disposal education. However, only about 52% of the respondents indicated that other community pharmacists were providing education. | Recommendations for disposal of non-controlled drugs: 4.2% flush down toilet/sink, 56% mix with undesirable substance and trash, 12% return to community pharmacy, 55% return to nearby fire or police station, 72.5% return on national drug take-back day, 28% other methods. | 80% said dropbox was not available in the city | not assessed | Lack of time, perceived patient disinterest, and absence of clear, unified guidelines on medication disposal as barriers to pharmacists providing disposal education. Confusion due to inconsistent recommendations by various organisations and limited pharmacist engagement in providing disposal education regularly. | Increase pharmacists' knowledge about medication disposal and providing strategies for enhancing patient-pharmacist interaction on this topic. The need for national guidelines and awareness programs mediated through healthcare professionals. |
